# Novel KIR2DS4:HLA-B*35 interaction predicts HLA-B*35 positive patient survival post hematopoietic stem cell transplant

**DOI:** 10.64898/2026.08.04.26358595

**Authors:** Stephen Gottschalk, Ying Li, Subodh Selukar, Alison M Kirk, Swati Naik, Daniel Fürst, Sophie Mannes, Sarah Flossdorf, Jan Beyersmann, Hubert Schezenmeier, Georg-Nikolaus Franke, the DRST Investigators, Paul G Thomas, Brandon M Triplett, Peter J Chockley

## Abstract

Haplo-identical hematopoietic cell transplantation (haploHCT) is an integral treatment paradigm for patients with leukemia. While overall survival (OS) post-haploHCT has steadily improved, relapse-free survival (RFS) remains relatively stagnant. Upon the discovery of killer immunoglobulin-like receptors (KIRs) on natural killer (NK) cells and their cognate human leukocyte antigen (HLA) ligands, algorithms have been developed to enhance graft versus leukemia effects. However, these algorithms fail to yield consistent predictions in patient outcomes.

We utilized a combination of *in silico* protein folding and interactions to determine KIR:HLA reactivity in conjunction with *in vitro* acoustic force microscopy to measure cell avidity (CA) as a readout for KIR signal strength. CA was determined using monoallelic HLA expressing K562 cell lines, monoallelic KIR Jurkat cells, and peripheral blood NK cells. We extended the CA results and performed standard cytotoxicity assays as well.

We discovered that HLA-B*35 interacts with KIR2DS4. We applied the newly discovered interaction to predict outcomes for HCT patients. Stratifying patients based on their HLA-B*35 positivity and donor KIR2DS4 status, we delineated a correlation to survival (P=0.061) when donors only had full-length KIR2DS4. Patients who received a haploHCT and NK cell addback from donors with only full-length KIR2DS4 had a significantly improved RFS (P=0.001) and OS (P=0.016) compared to truncated (KIR1D) and full-length KIR2DS4 donors. This was independently validated in a diverse 10/10 HLA matched European cohort with RFS (P=0.0255) and OS (P=0.0388).

Thus, the identified novel KIR2DS4:HLA-B*35 interaction axis predicts patient survival, in both haplo-identical and fully matched, HCT and highlights that our current understanding of the KIR:HLA interactome is incomplete and requires remapping for enhanced therapeutic applications.

## Introduction

The practice of hematopoietic cell transplantation (HCT) is nearing its seventh decade of clinical application^1^. Typically, the optimal donor is a fully HLA-matched sibling or unrelated donor, followed by a related haploidentical donor^2–4^. The use of haplo-identical donors for HCT (haploHCT) has been increasing in clinical practice^5^ due to the implementation of post-transplant cyclophosphamide^6^. HLA matching to avoid graft vs host disease (GvHD), driven mainly by αβT cells, has been the primary focus of donor selection, which could potentially limit graft versus leukemia (GvL) effects. In addition to αβT cells, natural killer (NK) cells, which are the first effector lymphocyte population to reconstitute in patients post-transplant^7–9^, have GvL activity^10^. NK cells specifically target cancer cells through an array of germline encoded receptors and their signals are integrated by NK cells to determine if lysis is to occur.

Killer immunoglobulin-like receptors (KIRs) expressed on NK cells mainly interact with HLAs^11,12^ and provide either activating or inhibitory signals to NK cells^11^. Recipient/donor KIR:HLA mismatch can drive GvL effects, resulting in enhanced survival^13–15^. Many selection and reactivity models based on KIR:HLA interactions have been proposed and implemented; however, none have been universally successful or consistent^16–18^. We posit that this failure is driven by an incomplete map of KIR:HLA interactions that drive NK cell reactivity to both healthy and malignant cells. Naturally occurring KIR variants add to the difficulty in creating KIR based selection algorithms; for example, KIR2DS4 exists in two isoforms, a full-length, functional, and an exon deleted, non-signaling and secreted, variant delineated as KIR1D^19,20^. Critically, KIR typing panels curated by EBMT and NMDP report both variants as “positive”^17^. Previous studies explored the role these variants may play in patient outcomes^19,21,22^, but there was no post-analysis of HLA haplotype to sub-categorize donor recipient pairs.

Herein, we describe a novel KIR2DS4:HLA-B*35 interaction that predicts patient survival in recipients of HCT, which lends credence for the need to revisit the previously defined categorization of KIR ligands and their downstream signaling effects.

## Materials and Methods

### *In silico* protein interactions

We took advantage of an *in silico* folding pipeline to assess the protein-protein interactions that drive the aggregated cell avidity and signal integrations between NK cells and tumor cells. AlphaFold3^23^ was used to predict protein interactions. Predicted aligned error (PAE) plots indicate accuracy of amino acid chain back bone position accuracy. Predicted template modeling (pTM) and the interface predicted template modeling (ipTM) are measures of overall subunit structure accuracy. pTM values above 0.5 indicate the structure may be similar to the true structure. ipTM values of ∼0.8 indicate a high confidence of complex subunit positioning.

### HLA-B*35 protein surface alignment

Additionally, we used AlphaFold3^23^ to predict the structure of the various HLA-B*35 mature protein sequences represented in our patient population from the Immuno Polymorphism Database^24^. These folded surfaces were then compared via the Protein Data Bank (PDB)^25^ to a known crystal structure of B*35:01 (4LNR^26^). The alignment scores (TM-score) of the protein surfaces were assessed along with the root mean squared distance (RMSD) of the carbon chain backbone with a TM-score of 1 being a perfect electron surface overlap and an RMSD of 0 a perfect bond alignment, respectively.

### Cell lines

HEK293T, K562, Jurkat, and NK92mi cell lines were grown according to the ATCC instructions. Primary NK cells were isolated and expanded as previously described^27^. Cells were routinely checked for mycoplasma contamination in house.

### Viral production and transduction

Gamma retroviral particles were produced as previously described^27^. Briefly, RD114 expressing HEK293T cells were transfected with 3.5μg of KIR2DS4.T2A.HALOtag containing plasmid DNA. Viral supernatants were isolated and filtered through 0.22micron filters and spun on to 0.125μg per well retronectin coated 24-well non-tissue coated plates for 90 minutes at 2000g. Viral supernatant was removed and cells were added at 500,000 cells per well and allowed to incubate overnight. Transduced cells were then removed, expanded, and checked for expression via flow cytometry and used for downstream analysis.

### Flow cytometry

We stained 250,000 of both primary NK cells and modified Jurkat and NK92mi cells with 1:200 dilution of anti- KIR2DS4-PE (Clone JJC11.6, Miltenyi Biotec, Bergisch Gladbach, Germany) antibody for detection on a BD FACSLyric flow cytometer and analyzed with FlowJo v10 (BD Biosciences, Franklin Lakes, NJ, USA).

### Cytotoxicity assay

K562 cancer cells were labeled with carboxyfluorescein succinimidyl ester (CFSE) 1:500 (Thermo Fisher Scientific, Waltham, MA, USA), and co-cultures with expanded NK cells were set at indicated ratios in 200 µL. After 24 h of co-incubation, 3,000 CountBright beads (Thermo Fisher Scientific, Waltham, MA, USA) were added to each well and assessed via flow cytometry for remaining CFSE-positive K562 cells. Samples were acquired until 100 beads were recorded. The percentage of remaining CFSE tumor cells compared with control tumor only wells was calculated according to the following equation: 100 - (CFSE Experimental/CFSE Control) x 100.

### Cell avidity measurements

We employed acoustic force microscopy, z-MOVI from Lumicks, LLC, (Amsterdam, The Netherlands), to directly interrogate cellular avidity between NK cells and target K562 cells. To ascertain specific HLA protein effects on NK cell binding we engineered a panel of mono-allelic HLA expressing cell lines. We employed known KIR:HLA combination controls, KIR2DS4:HLA-C*04 and KIR2DS4:HLA-C*07^28,29^. HLA-B*35:01, HLA-C*04:02, and HLA- C*07:01 molecules were expressed in K562 cells via lentiviral transduction. We isolated healthy donor peripheral blood NK cells that were subsequently KIR typed and used in cell avidity measurements against the mono-allelic HLA K562 cells. We compared the relative changes in cell avidity from the parental HLA null K562 cell lines and the mono-allelic HLA expressing sub lines. We performed antibody blockade of the KIR2DS4 receptor to specifically interrogate the role of this receptor in cellular avidity. Further, we created a mono-KIR2DS4 expressing Jurkat and NK92mi cell lines for cell avidity studies with the mono-allelic K562 HLA-B*35:01 cell line.

### Haploidentical and 10/10 HLA Matched HCT recipients

This retrospective study was reviewed and approved by a St Jude Institutional Review Board. HaploHCTs that were performed on or according to investigator-initiated protocols (NCT03849651, NCT01621477, NCT01807611, NCT02442660, NCT02790515, NCT02259348) between 2012, the year KIR typing was initiated at our institution^30,31^ and February 2024 were included in this analysis (**Supplementary Appendix Protocol Table**). We specifically analyzed patients receiving allogeneic HCT from a single KIR typed haploidentical donor for malignant disease (**Consort Diagram**, **Table 1**). Furthermore, with the same parameters as for our haploHCT patients, we analyzed 10/10 HLA matched HLA-B*35 positive subset of patients from the German Registry for Hematopoietic Stem Cell Transplantation and Cell Therapy (DRST)^19^ (**Table 3**). We have no access to patient or donor samples and is a clinically retrospective.

**Table 1:** Demographic, clinical, and donor characteristics of patients

|  | B*35+ Patients |  | B*35- Patients |  |
| --- | --- | --- | --- | --- |
|  | KIR2DS4 | KIR2DS4/1D | KIR2DS4 | KIR2DS4/1D |
| **Total** | 14 | 12 | 30 | 39 |
| **Male** | 8 | 6 | 16 | 21 |
| **Female** | 6 | 6 | 14 | 18 |
| **Age (yr)** |  |  |  |  |
| **Mean** | 11.2 | 10.1 | 9.4 | 11.4 |
| **Range** | (1.9-19.7)17.8 | (0.5-17.1)16.6 | (1.1-20.8)19.7 | (0.7-23.2)22.5 |
| **Patient CMV** |  |  |  |  |
| **Positive** | 11 | 10 | 22 | 31 |
| **Negative** | 3 | 2 | 8 | 8 |
| **Disease** |  |  |  |  |
| **AML** | 6 | 5 | 12 | 20 |
| **B-LL** | 4 | 5 | 16 | 12 |
| **T-ALL** | 2 | 1 | 1 | 2 |
| **Other** | 2 | 1 | 1 | 5 |
| **Disease Status** |  |  |  |  |
| **CR1** | 1 | 5 | 8 | 14 |
| **CR2** | 7 | 4 | 10 | 8 |
| **CR3** | 1 | 0 | 4 | 6 |
| **CR4** | 0 | 0 | 0 | 1 |
| **Relapse 1** | 3 | 1 | 4 | 5 |
| **Relapse 2** | 0 | 0 | 1 | 2 |
| **Relapse 5** | 0 | 0 | 0 | 1 |
| **REFR** | 1 | 1 | 3 | 2 |
| **PR** | 1 | 0 | 0 | 0 |
| **PIF** | 0 | 1 | 0 | 0 |
| **Conditioning Regimen** |  |  |  |  |
| **Myeloablative** | 3 | 0 | 3 | 1 |
| **Reduced Intensity** | 11 | 12 | 27 | 38 |
| **GvHD prophylaxis** |  |  |  |  |
| **Tac** | 6 | 2 | 9 | 7 |
| **Other** | 8 | 10 | 21 | 32 |
| **T cell Depletion** |  |  |  |  |
| **ATG or Alemtuzumab** | 4 | 4 | 8 | 14 |
| **Neither** | 10 | 8 | 22 | 25 |
| **Donor Type** |  |  |  |  |
| **Father** | 6 | 4 | 7 | 14 |
| **Mother** | 6 | 8 | 19 | 20 |
| **Grandparent** | 1 | 0 | 1 | 1 |
| **Sibling** | 1 | 0 | 3 | 4 |
| **Donor CMV** |  |  |  |  |
| **Positive** | 11 | 9 | 24 | 29 |
| **Negative** | 3 | 3 | 6 | 10 |
| **Unknown** | 0 | 0 | 0 | 0 |
| **Donor Sex** |  |  |  |  |
| **Male** | 6 | 4 | 7 | 17 |
| **Female** | 8 | 8 | 23 | 22 |
| **Donor Age (yr)** |  |  |  |  |
| **Mean** | 41.1 | 38.8 | 34.8 | 37.9 |
| **Range** | (23.6-55.2)31.6 | (25.9-53.2)27.3 | (18-49.7)31.6 | (20.6-54.6)34.0 |

Number of patients and donor age; CMV: Cytomegalovirus, AML: Acute Myeloid/Myelocytic/Megakaryoblastic Leukemia, B-ALL: B-cell lymphoblastic leukemia, T-ALL: T cell acute lymphoblastic leukemia, CR1: first complete remission CR $\geq 2$ , second or later CR, REFR: Refractory, PR: Partial Response, PIF: Primary engraftment failure, Tac: Tacrolimus, ATG: Anti-thymocyte globulin

### Statistical analysis

We analyzed patient data descriptively and with survival analysis methods. We estimated the overall survival (OS, events: all-cause mortality) and relapse-free survival (RFS, events: relapse or all-cause mortality) probabilities using the Kaplan-Meier method with an index date of 100 days post-haploHCT using the landmark approach^32^. For OS, event-free patients were censored at last contact, and for RFS, event-free patients were censored at last contact or 5 years post-haploHCT, whichever came first. Primary analyses evaluated the B*35+ cohort and secondary analyses evaluated the B*35- cohort, and we repeated analyses in subgroups of patients who received protocol-specified donor NK cells 6 days post-transplant (“HAPNK” subgroups). In the main text, we report results from analyses using the difference in restricted mean survival times (RMST) with a time horizon of 5 years post-haploHCT (equal to 4.73 years after 100 days post-haploHCT). In the **Supplementary Appendix**, we detail sensitivity analyses using other time horizons and adjusting for potential confounders (disease status and CMV status) and supplementary analyses with alternative approaches. Inferential results are intended to be hypothesis generating and are not adjusted for multiple comparisons. DRST cohort patients were assessed using log-rank test.

## Results

### Determination of novel KIR:HLA interaction

Based on donor variability in our studies using allogeneic NK cells in various pre-clinical tumor models^27,33^, we found a pattern that implicated a novel KIR:HLA interaction. KIR2DS4 and HLA-B*35 status appeared to be a predictor of improved cytolytic NK cell interactions with cancer cells and their antitumor activity. *In silico* folding and interaction analyses using Alphafold3 (**Figure 1A,B**) indicated a potential interaction between KIR2DS4 and HLA-B*35. This result was compared to two known KIR2DS4 binders, HLA-C*04:01 and C*07:02 and one non-binder HLA-A*01:01 (**Figure S1A-C**). The predicted interactivity scoring was similar to that of KIR2DS4 and HLA-B*35.. To uniquely investigate the specific KIR2DS4:HLA-B*35 binding axis, we engineered a T cell receptor (TCR) and KIR negative Jurkat cell line to express KIR2DS4 (**Figure 1C**). These cells bound monoallelic HLA- B*35 expressing K562 cells to an exceptionally high degree (**Figure 1D**). Conversely, we observed parental Jurkat cells binding equally to both K562 and K562 B*35 cells (**Figure 1E**). This was repeated with NK92mi cells (**Figure 1F,G,H**), respectively.

**Figure 1:**
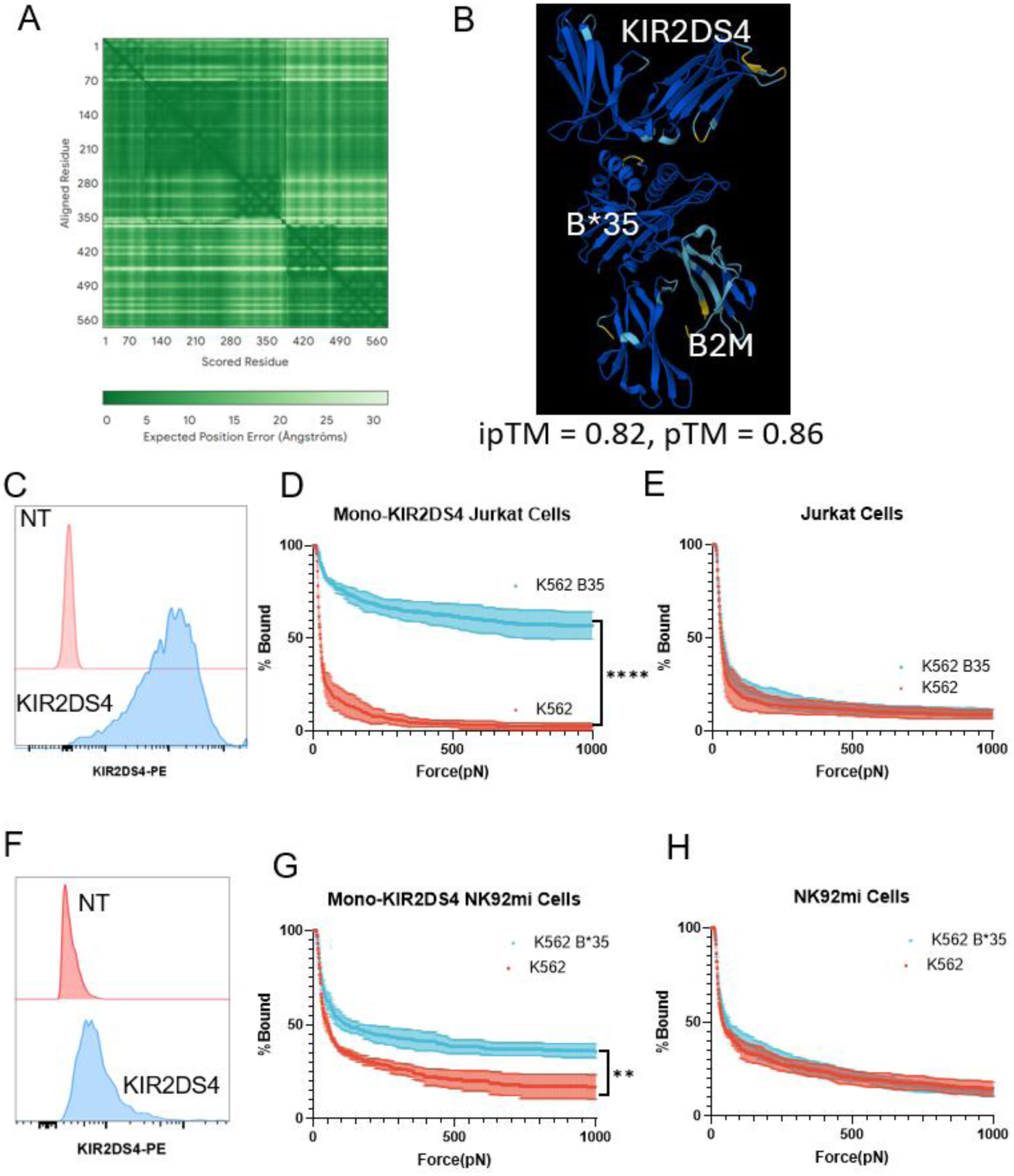
Novel KIR:HLA interaction partners KIR2DS4 and HLA B*35. (A) *In Silico* Alphafold3 prediction indicating high fold confidence as shown by the predicted position alignment error (PAE) plot. (B) 3-D rendering of spatial location of KIR2DS4 and peptide loaded HLA-B*35:01 in complex with beta-2 microglobulin. Indicated values of ipTM, 0.82 and pTM, 0.86. (C) Mono-KIR2DS4 expressing Jurkat cells labeled with anti-KIR2DS4 flow cytometry histogram plot (D) Mono-KIR2DS4 expressing Jurkat cells in cell avidity assays with K562 Parental and mono-allelic HLA- B*35:01 expressing K562 cells. AUC analysis n=3 independent experiments, paired t-Test, p<0.0001 **** (E) Jurkat cells in cell avidity assays with K562 Parental and mono-allelic HLA-B*35:01 expressing K562 cells. (F) Mono-KIR2DS4 expressing NK92mi cells labeled with anti-KIR2DS4 flow cytometry histogram plot (G) Mono-KIR2DS4 expressing NK92mi cells in cell avidity assays with K562 Parental and mono-allelic HLA-B*35:01 expressing K562 cells. AUC analysis n=3 independent experiments, paired t-Test, p<0.01 ** (H) NK92mi cells in cell avidity assays with K562 Parental and mono-allelic HLA-B*35:01 expressing K562 cells.

Applying the same experimental parameters, we performed functional studies with peripheral blood NK cells from an array of healthy donors (**Figure 2A**). Donors with KIR2DS4 and not KIR2DS1 were analyzed via flowcytometry (**Figure 2B**).. To directly measure the influence of single HLA molecules on cell avidity, a panel of mono-allelic K562 cell lines that only express the indicated HLA molecules (**Figure 2C**) were assessed for cell binding to each donor. To illustrate this assay, greater binding of donor 3 NK cells to HLA-B*35-expressing K562 cells vs the parental line is shown in **Figure S2C,D**, indicating enhanced activation and stronger synapse formation. Additionally, we blocked this interaction with an KIR2DS4 antibody and found that the cell avidity was only blocked when K562 cells expressed HLA-B*35 (**Figure S2D-H**). We sought to investigate the cytotoxic profiles of these various donor NK cells and observed no difference with KIR1D Donors (**Figure 2D**). This was juxtaposed by KIR2DS4 positive NK cells targeting mono-allelic K562 B*35 cells more (**Figure 1E**). Thus, our data demonstrates specific interaction between these donor NK cells and cell lines, which strongly argues against alternative KIR:HLA interactions contributing to enhanced binding of KIR2DS4-positive primary NK cells to HLA-B*35 expressing cancer cells.

**Figure 2:**
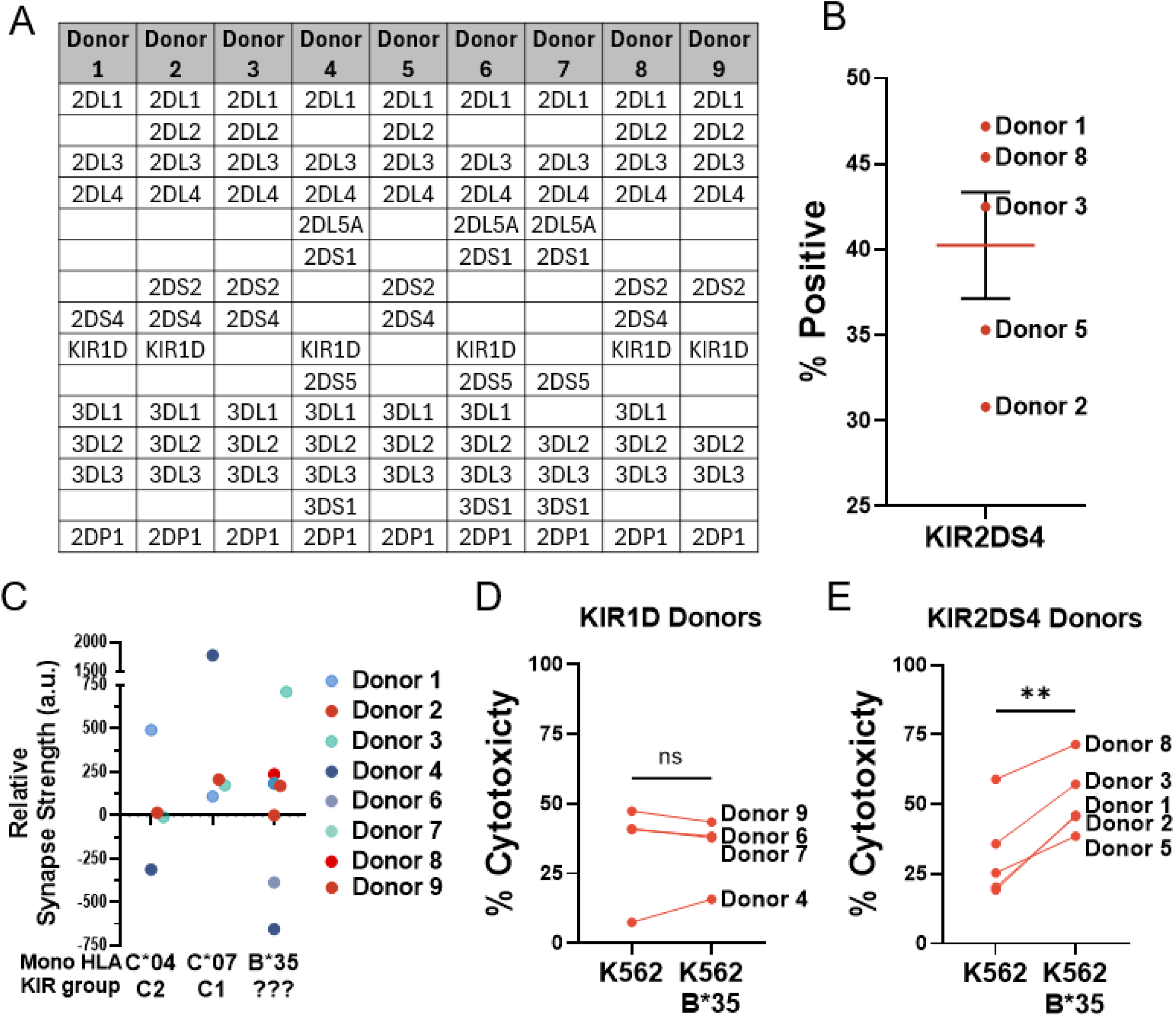
KIR2DS4 binding HLA-B*35 increases cell avidity and cytotoxicity in primary NK cells. (A) Genotypic KIR typing performed on 9 healthy donor peripheral blood NK cells. (B) Percent positive NK ells for KIR2DS4 as determined by flow cytometry or indicated donors. (C) Functional assessment of the donors towards indication mono-allelic HLA expressing K562 cells. The change in relative binding is the difference between areas under the curves of each cell avidity measurement throughout a force ramp of 0-1000pN. The HLA null parental K562 served as the basal binding level of each donor NK cell. Positive values indicate increased binding strength and higher activation levels. Published KIR reactivity groups are listed for C*04:02 as C2 and C*07:01 as C1 each n=4 and B835:01 n=8. (D) KIR1D donor 24-hour cytotoxicity assay with K562 Parental and mono-allelic HLA-B*35:01 expressing K562 cells, n=4 paired t-Test, ns (not significant) (E) KIR2DS4 donor 24-hour cytotoxicity assay with K562 Parental and mono-allelic HLA-B*35:01 expressing K562 cells, n=5 paired t-Test, p<0.01 **

### Patient stratification and clinical evaluation

Due to the age and variability in technologies utilized for haplotyping patients, we grouped all HLA-B*35 molecules together, i.e. HLA-B*35:01 through HLA-B*35:43 as these were expressed in our patient population. We ensured HLA-B*35 protein surface similarity using AlphaFold3 and Protein Data Bank (PDB) alignments (**Figure S3A-C**). Donors were stratified based on their KIR2DS4 genotype profiles. Donors expressing full length and functional KIR2DS4 from both alleles are denoted as “KIR2DS4”, and mixed expressing donors with both the full length and the deleted non-functional variant of KIR2DS4 as “KIR2DS4/1D”. Patients were limited to those that had been treated with a haploHCT (**Consort diagram**). All patients were treated for hematological malignancies, predominantly acute myeloid leukemia (AML) and B cell acute lymphoblastic leukemia (B-ALL). Complete patient characteristics of each group are listed in **Table 1**.

To aid in comparison to other published reports^17^ regarding the presence of KIR2DS4, we delineated our entire donor population according to full length and deleted exon isoforms and observed the following haplotype frequency: purely full length, 25.1%; mixed full length and truncated, 26.25%; truncated only, 45.17%; and 3.47% fully absent. Thus, the KIR2DS4 or KIR1D positive rate was 96.53%, and only 51.35% of the population had functional KIR2DS4 proteins (**Figure S4**).

We asked if increased functional donor KIR2DS4 levels resulted in any survival advantage of patients that were HLA-B*35 positive. We analyzed 26 patients in the HLA-B*35 positive cohort with a median time from haploHCT to death or censoring of 3.63 years. There was a total of 10 deaths and 11 RFS events. We observed that KIR2DS4 donors resulted in enhanced OS (P=0.061) (**Figure 3A**, **Table 2**) and RFS (P=0.055) (**Figure 3B**, **Table 2**) of patients compared to KIR2DS4/1D donors. Next, we sought to ensure that this effect is specific to KIR2DS4:HLA-B*35 interactions. We selected HLA-B*35 negative patients and assessed them for survival in the same donor groupings. In the HLA-B*35 negative cohort, we analyzed 69 patients with a median time from haploHCT to death or censoring of 3.97 years, with a total of 22 deaths and 25 RFS events. Intriguingly, we found an inverse relationship where KIR2DS4/1D donors trended with better OS (P=0.046) (**Figure S5**, **Table 2**). As expected, when using shorter time horizons, strengths of association are lower (**Table S1**). In sensitivity analyses, we did not find evidence that our results are confounded by disease or CMV status (**Tables S2-6**). We also repeated the 5-year RMST analysis without the landmark approach (OS and RFS measured from haploHCT not 100 days post-haploHCT), and we found that directions of association were identical, but strengths of association were lower, as expected (**Table S7**). Additionally, we analyzed purely KIR1D donors in both patient HLA-B*35 positive (**Figure S6A**) and negative (**Figure S6B**) populations. In HLA-B*35 positive patients and we found a mildly reduced survival compared to the KIR2DS4 donor group and in the HLA-B*35 negative setting a similar result to the KIR2DS4 donor group.

**Figure 3:**
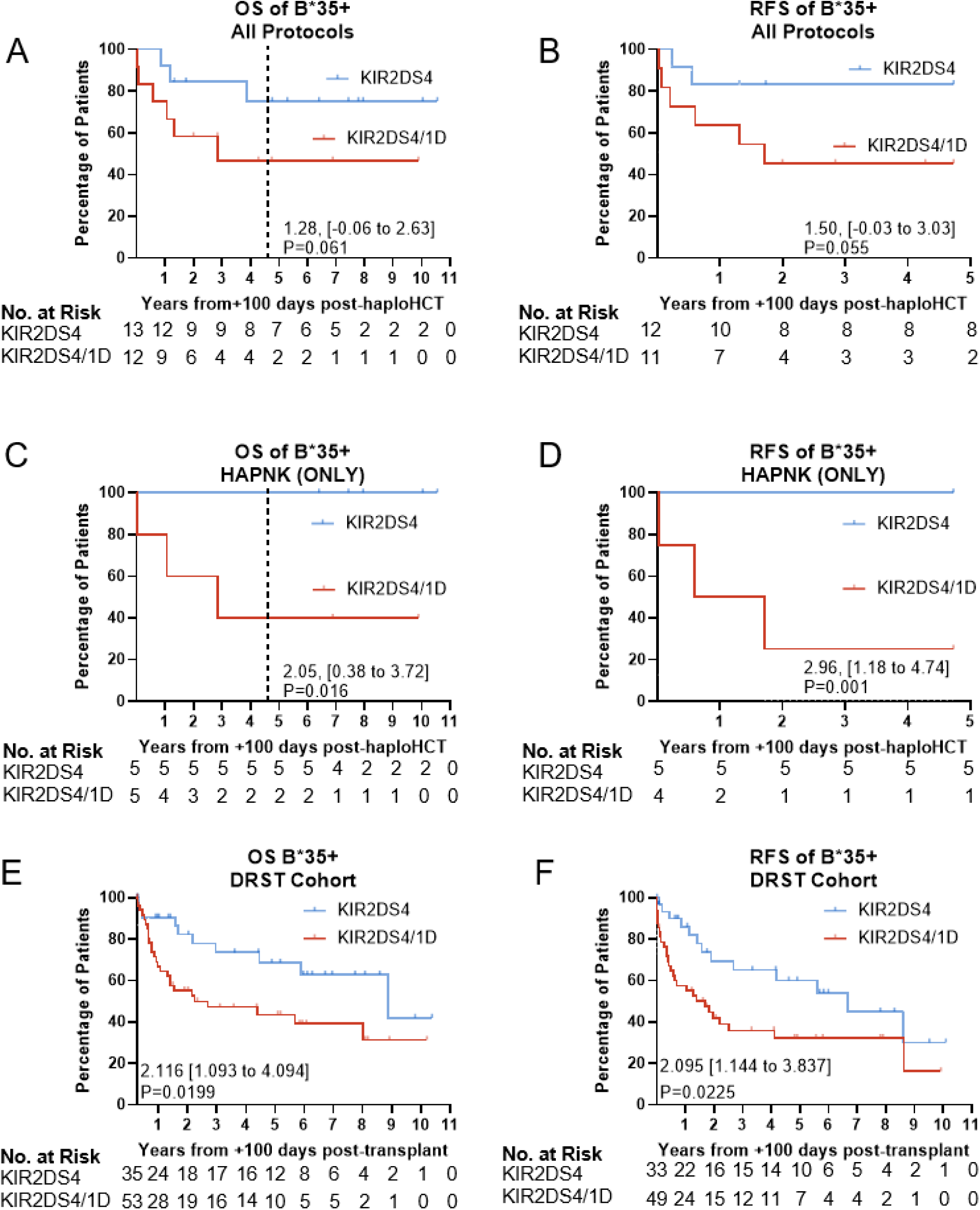
HCT outcomes of patients stratified on donor KIR2DS4 typing. Event-free patients at day 100 post-haploHCT were separated based on their HLA-B*35 status and then subset by KIR2DS4 full length only (KIR2DS4) or full length with deleted variant (KIR2DS4/1D). Difference in 5-year RMST (years) is indicated with change from reference KIR2DS4/1D indicated with 95% confidence interval upper and lower bounds in brackets and two-sided Wald-based p-value. Dashed lines indicate 4.73 years. The number of patients at risk over the years is indicated below each graph. (A) HLA-B*35 positive patient overall survival in all protocols. (B) HLA-B*35 positive patient relapse free survival in all protocols. (C) Specific clinical protocol HAPNK^34^ on which patients received a single infusion of their haploHCT donors peripheral blood NK cells on day 6 post-transplant, overall survival. (D) HAPNK patients relapse free survival. (E) Overall survival from independent validation cohort DRST of HLA-B*35 positive patients. Hazard ratios and two-sided p-value from Log-Rank test shown. (F) Relapse free survival from independent validation cohort DRST of HLA-B*35 positive patients. Hazard ratios and two-sided p-value from Log-Rank test shown.

**Table 2:** 5-Year restricted mean survival times.

| Cohort | Endpoint | KIR2DS4/1D 5-year RMST (years) | KIR2DS4 5-year RMST (years) | Difference in 5-year RMST (years, reference: KIR2DS4/1D) | 95% LB (years) | 95% UB (years) | P-Value |
| --- | --- | --- | --- | --- | --- | --- | --- |
| **B\*35+** | OS | 2.79 | 4.07 | 1.28 | -0.06 | 2.63 | 0.061 |
|  | RFS | 2.51 | 4.00 | 1.50 | -0.03 | 3.03 | 0.055 |
|  | OS, HAPNK | 2.68 | 4.73 | 2.05 | 0.38 | 3.72 | 0.016 |
|  | RFS, HAPNK | 1.77 | 4.73 | 2.96 | 1.18 | 4.74 | 0.001 |
| **B\*35-** | OS | 4.10 | 3.29 | -0.82 | -1.62 | -0.02 | 0.046 |
|  | RFS | 3.95 | 2.98 | -0.97 | -1.86 | -0.07 | 0.034 |
|  | OS, HAPNK | 4.13 | 3.30 | -0.82 | -2.10 | 0.45 | 0.205 |
|  | RFS, HAPNK | 4.09 | 3.00 | -1.09 | -2.55 | 0.37 | 0.142 |

Differences in 5-year restricted mean survival times (RMSTs) in years for overall survival (OS) and relapse-free survival (RFS) endpoints, B\*35+ and B\*35- cohorts, and the corresponding HAPNK subgroups. Lower Bound (LB) of 95% confidence interval, Upper Bound (UB) of 95% confidence interval, specific NK cell addback protocol (HAPNK).

**Table 3:** Demographic, clinical, and donor characteristics of +100 Days post-HCT DRST cohort patients.

|  | **B\*35+ Patients** |  |
| --- | --- | --- |
|  | **KIR2DS4** | **KIR2DS4/1D** |
| **Total** | 35 | 53 |
| **Age (yr)** |  |  |
| **Mean** | 47.2 | 49.2 |
| **Range** | (8-68)60 | (6-71)65 |
| **Patient CMV** |  |  |
| **Positive** | 20 | 30 |
| **Negative** | 14 | 22 |
| **Unknown** | 1 | 1 |
| **Disease** |  |  |
| **AL** | 4 | 0 |
| **ALL** | 5 | 8 |
| **AML** | 13 | 23 |
| **CLL** | 4 | 4 |
| **CML** | 0 | 1 |
| **NHL** | 1 | 1 |
| **Lymphoma** | 6 | 10 |
| **MM** | 1 | 5 |
| **Other** | 1 | 2 |
| **Disease Status** |  |  |
| **Ealy** | 18 | 18 |
| **Intermediate** | 10 | 22 |
| **Advanced** | 7 | 13 |
| **Conditioning Regimen** |  |  |
| **Myeloablative** | 25 | 31 |
| **Reduced Intensity** | 10 | 22 |
| **T cell Depletion** |  |  |
| **ATG** | 32 | 43 |
| **Neither** | 1 | 12 |
| **Unknown** | 2 | 8 |
| **Donor CMV** |  |  |
| **Positive** | 15 | 20 |
| **Negative** | 19 | 32 |
| **Unknown** | 1 | 1 |
| **Donor Age (year range)** |  |  |
| **18-30** | 12 | 16 |
| **31-45** | 16 | 26 |
| **46-60** | 6 | 8 |
| **missing** | 1 | 3 |

Number of patients and donor age; CMV: Cytomegalovirus, AL: Acute Leukemia, ALL: Acute Lymphoblastic Leukemia, AML: Acute Myeloid Leukemia, CLL: Chronic Lymphocytic Leukemia, NHL: non-Hodgkins Leukemia, MM: Multiple Myeloma, ATG: Anti-thymocyte globulin

To further elucidate the role of KIR2DS4 on NK cells, we considered a subset of the total patients that received an infusion of donor NK cells on day 6 post-transplant (HAPNK protocol)^34^. Strikingly, no deaths occurred among HLA-B*35 patients receiving KIR2DS4 donor NK cells, resulting in longer OS than corresponding patients who received KIR2DS4/1D donor NK cells (P=0.016) (**Figure 3C**, **Table 2**). Furthermore, no relapses were observed in the KIR2DS4 donor group, resulting in a significantly longer RFS than in the KIR2DS4/1D donor group, (P=0.001) (**Figure 3D**, **Table 2**).

We extended these findings to an independent, 10/10 HLA matched, validation cohort from the DRST. Patient grouping profiles are detailed in (**Table 3**) and are similar in all parameters. Patients were stratified as before and again we find that 10/10 HLA matched patients had increased overall survival (p=0.026) (**Figure 3E**) and relapse-free survival (p=0.0225) (**Figure 3F**).

## Discussion

We have discovered a novel interaction between KIR2DS4 and HLA-B*35 molecules using *in silico* and *in vitro* assays. This interaction predicted OS and RFS across haploHCTs conducted at our center since 2012, most strikingly in the immune effector addback setting with NK cells. Furthermore, the validation cohort of 10/10 HLA matched patients with a range of diseases and ages confirmed the same significant pattern of enhanced patient outcomes. These differences were evident despite differences in patient age, underlying leukemia diagnosis, disease status at transplant, and conditioning regimen used, highlighting the cross-cutting impact of this novel KIR:HLA interaction. Our understanding of KIR:HLA interactions is still limited in scope in large part due to the sheer number of possible combinations. The two genomic loci represent the most diverse loci across humans and are responsible for numerous critical immunological functions^35^. This diversity allows for our resilience against foreign pathogens, cancer immunosurveillance, and fetal development.

Numerous clinical trials have explored various KIR-based outcome predictions in HCT, but in the end have not yielded comparable or conclusive results to indicate an optimal selection process thus far^18^. Based on recent publications, considering KIR:HLA interactions in the donor selection process are not considered to be clinically meaningful^17^, and there is an extensive body of work showing limited correlation with transplant outcomes^18^. We posit that this is due to an outdated and incomplete KIR reactivity profile since recombinant Fc fusion protein binding has been predominately used to determine KIR:HLA interactions^36^. This allows only for the detection of high affinity interactions, while failing to predict aggregate binding that occurs between two cells that express thousands of molecules on their cell surface. Supporting this notion is a recent study identifying biparatopic antibodies in which avidity measurements predicted efficacy *in vivo*^37^.

Specifically, what is not interrogated with the use of Fc fusion protein binding is the ability for an NK cell to integrate weaker signals that can lead to a cascading signaling effect and result in activation or inhibition. Productive interactions between NK and target cells result in the formation of an immunological synapse, which is directly controlled by the relative activation level that the NK cell senses^38^. Methods to easily and reproducibly interrogate the strength of cellular synapses have only recently become available^39^. We have employed an acoustic force microscope, z-MOVI, to measure the cell avidity on the piconewton scale^33^, and directly assessed cell-cell interactivity in a controlled manner whereby only single HLA molecules were contributing to NK cell binding. We confirmed the specificity of interaction by utilizing a mono-KIR2DS4 expressing cell lines as well as antibody blockade of the KIR2DS4 receptor on peripheral blood NK cells.

We validated our findings in a 10/10 HLA matched patient cohort from the DSRT indicating that the phenotype is not restricted to the haploHCT setting. In addition, we explored other registries, including NMDP and EBMT databases, but found that these do not delineate patients and donors for full length, functional, and exon deleted, non-signaling, variant of KIR2DS4 (personal communications). Recent studies have found that KIR2DS4 was among a signature for donor profiles that resulted in reduced relapse rates post-HCT^17^. This is unsurprising as KIR2DS4 was reported as “positive” in a near uniform manner in the analyzed donor pool. Based on our findings, we suggest an added layer of refinement in this respect to aid in parsing between donors with functional (full length) or non-signaling (truncated) KIR2DS4 variants as our work, as well as previous studies^19,21,22^, reveal the import of this distinction.

One limitation of our study is that the pediatric sample size is relatively low, limiting statistical power and certain statistical analysis methods. We conducted multiple sensitivity and supplementary analyses, and these largely corroborated the primary 5-year RMST results, confirming that directions of association are not sensitive to modeling choices, but strengths of association differ, which we attributed primarily to the sample size and numbers of events (**Supplementary Appendix**). We conducted confounder-adjusted analyses, which did not suggest evidence for sensitivity to confounding factors in this data set. We focused on outcomes after day 100 post-haploHCT since early events are, in general, treatment related. In our data set, events before day +100 were similar between groups, and the directions of association remained the same in supplementary analyses that included these early events.

One intriguing facet that we observed was that patients that are HLA-B*35 negative benefited with a mixed expression of KIR2DS4 and KIR1D HCT donor. We hypothesize that the secreted KIR1D is not entirely inert to NK cell reactivity and may mask other HLA proteins as a decoy and thus augment NK reactivity by cloaking either activating or inhibitory KIRs. Further, adding to the nuanced biology of KIR2DS4, it is the only activating KIR on the A locus and may play a role in modulating or tuning the B locus activating KIRs. Further support for the KIR1D decoy hypothesis comes from the observation that patients receiving transplants from donors with a KIR A/A haplotype and KIR1D had improved survival^19^. However, no subsequent patient HLA stratification was performed. In our dataset, fully KIR1D donors displayed a slightly reduced HLA-B*35 positive patient survival compared to full KIR2DS4 donors and a similar survival rate in HLA-B*35 negative patients, both being reduced compared to the mixed KIR2DS4/1D donors. We are actively designing truncated KIR1D variants for testing in our binding and killing assays to see which HLA molecules may be impacted by soluble KIR1D protein binding.

In conclusion, we have discovered a novel interaction between KIR2DS4 and HLB-B*35 that resulted in enhanced RFS and OS post-HCT. The ability to select the optimal donor is paramount to increase graft versus leukemia activity and reduce adverse effects post-transplant. Our work indicates that our current understanding of the KIR:HLA interactome is incomplete and requires remapping utilizing modern techniques and tools to aid in donor selection and, ultimately, to enhance patient survival.

## Data Availability

All data produced in the present study are available upon reasonable request to the corresponding author.

## Acknowledgements

We thank the patients, their caregivers and families; the dedicated nurses and physicians who cared for them; and the staff members involved in data collection and analyses. We would like to especially thank the HLA and KIR typing support from Paula Arnold and the HLA lab in the Department of Pathology at St Jude Children’s Research Hospital. Additionally, the members of the DRST for curating and supplying the additional patient cohort listed in the appendix. This study was supported by the National Institutes of Health (NIH)/National Cancer Institute (NCI) grant P30CA021765, NIH/NCI K22CA292567, The Edward P. Evans Foundation, and the American Lebanese Syrian Associated Charities (ALSAC). The content is solely the responsibility of the authors and does not necessarily represent the official views of the NIH.

## Author contributions

SG, SS, and PJC wrote original manuscript. SG, YL, SS, AMK, DF, SM, SF, JB, HS, GNK, PGT, BMT, PJC performed experimentation and/or supported research for data generation. SS and PJC performed statistical analyses. Conceptualization and study oversight PJC.

## Consort Diagram

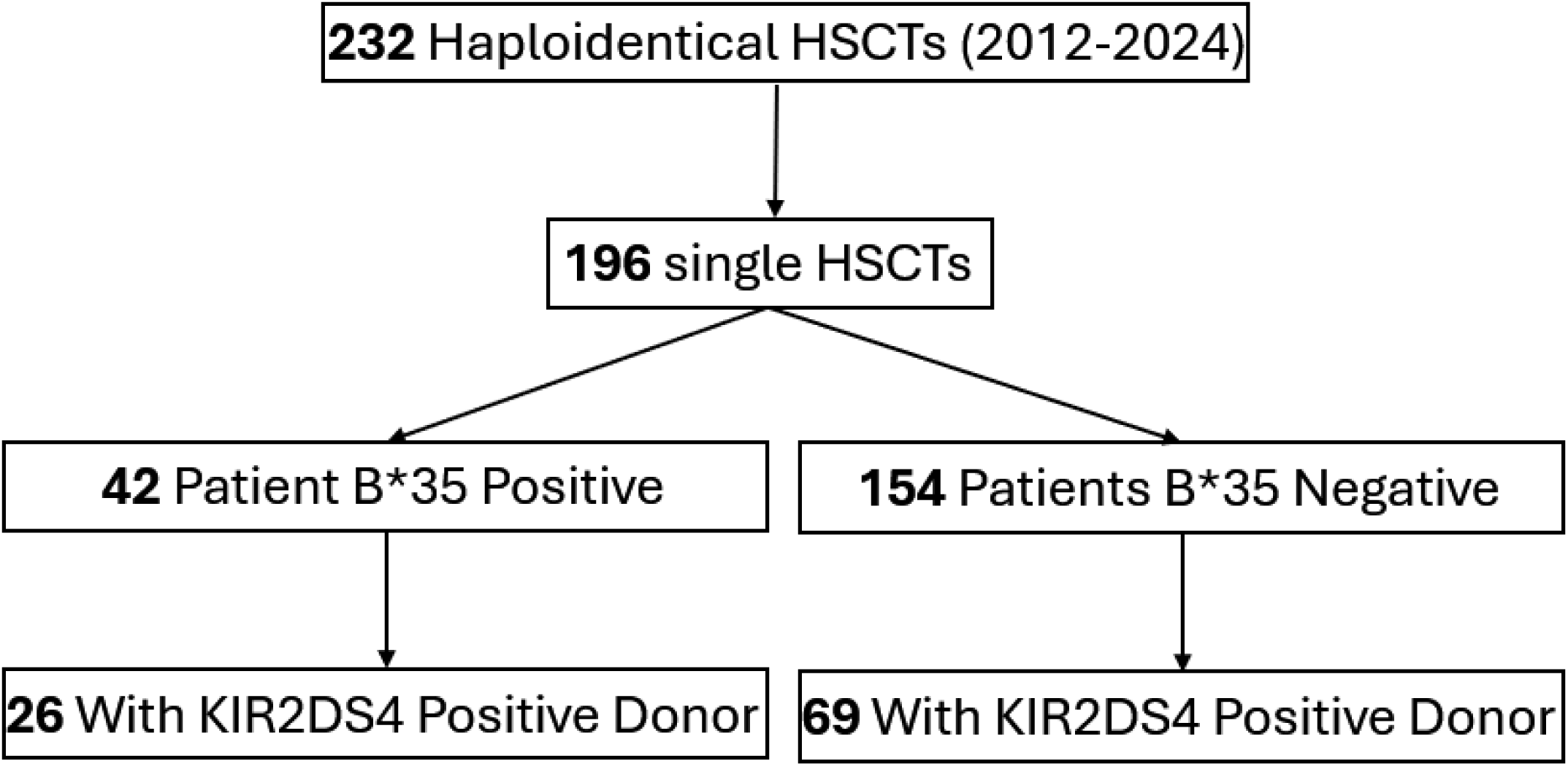

## Supplementary Materials

### Cell avidity protocol

K562 (ATCC # CCL-243) myeloblastic leukemia is a common NK cells target and is HLA null. Briefly, K562 cells are loaded into a concanavalin A coated z-MOVI chip and allowed to adhere. Human NK or Jurkat cells (Sigma Aldrich # SCC635) are labeled with cell trace far red (Thermo Scientific #34564) per manufacturer’s instructions. NK cells are then loaded into the chip and allowed to interact with the K562 monolayer for 5 minutes, 2 minutes for Jurkat cells, before being subjected to an acoustic piconewton (pN) force ramp over 2.5 minutes from 0pN to1000pN. Individual cells are observed via fluorescence microscopy as they release from the K562 cells on a single cell resolution level with specific pN forces recorded. In some cases, NK cells were incubated with KIR2DS4 antibodies (Miltenyi Biotec JJC11.6 PE-Conjugated) for 30 minutes prior to loading on to the avidity chip.

### Monoallelic KIR2DS4-expressing cell line

The full-length coding sequence for KIR2DS4*0010101 was obtained from the IPD-IMGT/HLA database^24^, then synthesized with a HALO tag on the C-terminus and cloned into an gamma retro viral vector (Genscript). Virus was packaged with 293vec-RD114 (Biovec pharma), isolated virus was spun onto retronectin (Takeda #T100B) coated plates for 1.5 hours at 2000g. Jurkat cells were incubated on virus coated plates overnight and culture per manufacturer’s instructions. Transduction efficiency was determined via flow cytometry with the aforementioned anti-KIR2DS4-PE.

### Monoallelic HLA-expressing cell lines

The full-length coding sequences for HLA alleles B*35:01, C*04:02, and C*07:01 were obtained from the IPD-IMGT/HLA database^24^, then synthesized and cloned via restriction sites (Genscript) into lentiviral vector pLVX- EF1α-IRES-Puro (Takara 631253). Lentivirus was packaged by transfecting HEK 293T cells (ATCC CRL-3216) with the HLA-containing lentiviral vector, psPAX2 packaging plasmid (a kind gift from Didier Trono via Addgene; Addgene plasmid #12260), and the pMD2.G envelope plasmid (a kind gift from Didier Trono via Addgene; Addgene plasmid #12259) at a ratio of 4:3:1. Viral supernatant was harvested at 24 and 48 hours post-transfection and filtered through a 0.45 µm SFCA syringe filter (ThermoFisher Scientific 723-9945), then concentrated using Lenti-X Concentrator (Takara 631232). Single-use aliquots of concentrated lentivirus in PBS were frozen at -80 °C until use. K562 cells (ATCC CCL-243) were transduced with concentrated lentivirus, and 72 hours post-transduction, puromycin (Sigma-Aldrich P9620) was added at 2 μg/mL to select for transduced cells. After one week of antibiotic selection, transduction and surface HLA expression was confirmed via flow cytometry using a pan-HLA class I antibody (PE-conjugated, Biolegend 311406, clone W6/32).

### Detailed patient data survival analysis methods

In this section, we detail the analyses conducted for the survival analysis of patient data. This is a retrospective data analysis, and there was no prespecified analysis plan.

The primary analysis population is the B*35+ cohort, and the B*35- cohort is a secondary analysis population. We also explored the respective subgroups of patients who received protocol-specified NK cell addback (denoted as “HAPNK”) for a total of 4 analysis populations.

The primary endpoint is overall survival (OS) measured from day 100 post-haploHCT to all-cause mortality. The secondary endpoint is relapse-free survival (RFS) measured from day 100 post-haploHCT to relapse or all-cause mortality. For OS, event-free patients were censored at last contact. For RFS, event-free patients were censored at last contact or 5 years post-haploHCT, whichever came first. This was chosen because of anticipated lower quality of relapse reporting after 5 years post-haploHCT, while survival status could be obtained with longer follow-up. We repeated analyses for both endpoints and all 4 analysis populations.

Consistent with the landmark approach^32^, patients with events or censoring before 100 days post-haploHCT were excluded from respective analyses. The study design excluded patients with unavailable KIR2DS4 grouping. The analysis data did not include any missing data (beyond censored data addressed by the analysis choices).

Each analysis compares the two KIR2DS4 groups for the respective endpoint (reference group: KIR2DS4/1D). For descriptive purposes, we visualized Kaplan-Meier (KM) curves for each analysis. The primary statistic for all analyses is the restricted mean survival time (RMST) at 5 years post-haploHCT, consistent with the RFS follow-up and highest quality evidence. In addition to group-specific RMSTs, we computed the difference in RMSTs, the Wald-based 95% confidence interval for the difference, and the two-sided Wald-based p-value. These calculations used the survRM2 R package’s rmst2 function.

We conducted two sensitivity analyses:

1. We repeated the analyses while varying time horizons. We used the rmst2 default time horizon, approximating the longest available follow-up time (labeled as “max” in tables below), and we used shorter times of 1, 2, 3 and 4 years post-haploHCT. We provide the p-values by time horizon below.
2. We repeated the 5-year RMST analysis adjusting for potential confounders of disease status (categorized as complete remission [CR] or other) and CMV status (categorized as positive, negative or unknown). We report the adjusted difference in RMSTs, the Wald-based 95% confidence interval for the adjusted difference, and the two-sided Wald-based p-value.

We conducted supplementary analyses using alternative statistics:

1. We analyzed the 5-year KM estimates similar to the 5-year RMSTs: we computed the difference in 5-year KM estimates, the Wald-based 95% confidence interval for the difference, and the two-sided Wald-based p-value. We used the survival R package for these calculations.
2. We analyzed the Cox proportional hazards model. Due to limitations in the sample size and numbers of events, we used conventional Cox models and Firth-penalized Cox models^40^ (“firthCox” models). We used the survival and coxphf R packages for these calculations (for the coxphf functions we adjusted the maximum iterations to 1000 from the default 50).

a. Conventional Cox model: We fit conventional Cox models for the B*35 positive and negative cohorts for both OS and RFS endpoints. For the B*35+ cohort, the model could not be fit for the HAPNK subgroup, but it could be fit for the B*35- cohort subgroup. Multivariable Cox models could not be fit using conventional Cox models. We report unadjusted hazard ratios with Wald-based 95% confidence intervals. We computed two-sided p-values in two ways: likelihood ratio test (LRT) and score test (equivalent to log-rank test).
b. firthCox model: We fit univariable firthCox models for both OS and RFS for all 4 analysis populations. We also fit corresponding multivariable firthCox models. From this, we report unadjusted and adjusted hazard ratios with respective penalized LRT (pLRT)-based 95% confidence intervals, and we compute two-sided pLRT-based p-values.

As a separate supplementary analysis, we also analyzed OS* and RFS* defined, respectively, as OS and RFS measured from date of haploHCT instead of day 100 post-haploHCT (i.e., not using the landmark approach). For this, we repeated the 5-year RMST analysis with OS* and RFS* for all 4 analysis populations. This analysis included all patients and did not exclude patients with events or censoring before day 100 post-haploHCT.

#### Results from sensitivity and supplementary analyses

In this section, we provide the results from the sensitivity and supplementary analyses. Except where otherwise specified, results use the 100-day landmark analysis populations.

### Cox Modeling

Based on the plateau regions of the Kaplan-Meier curves and limited events in the KIR2DS4 group of the B*35+ cohort, we believe that these data may not be well-represented by modeling that assumes proportional hazards. For this reason, we elected to use Cox models only as supplementary analyses.

We encountered model fitting issues due to the limited numbers of events in the HAPNK subgroup of the B*35+ cohort. Cox model results tend to show somewhat weaker evidence of association for the B*35+ group (relative to the 5-year RMST analysis), while they show stronger evidence of association for the B*35- cohort. LRT-based and score test-based p-values are similar to each other.

**Figure S1:**
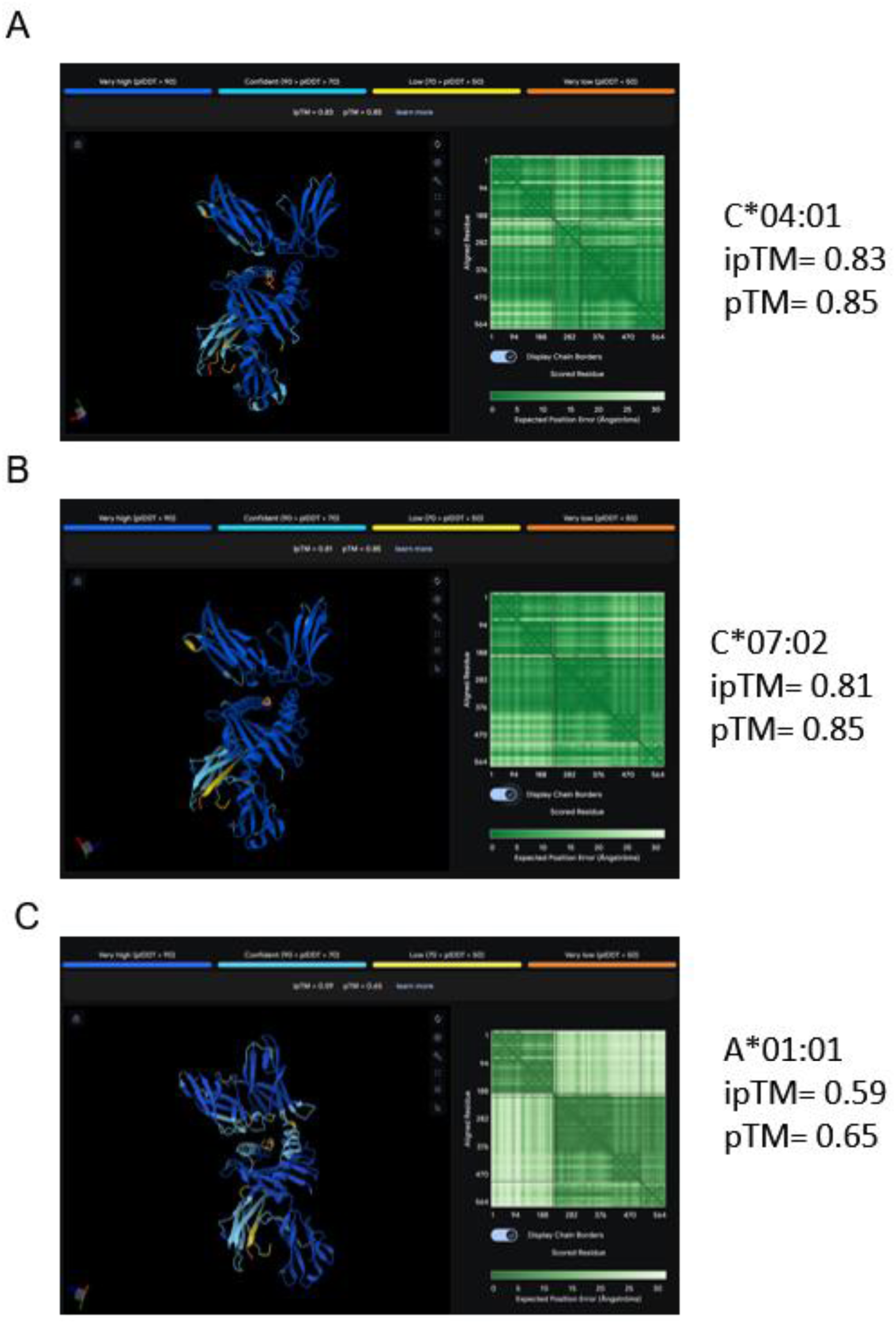
Comparison of control HLA protein interactions with KIR2DS4. *In Silico* Alphafold3 prediction indicating fold confidence as shown by the predicted position alignment error (PAE) plot and ipTM and pTM values. (A) HLA-C*04:01 known binder of KIR2DS4 (B) HLA-C*07:02 known binder of KIR2DS4 (C) HLA-A*01:01 known non-binder of KIR2DS4

**Figure S2:**
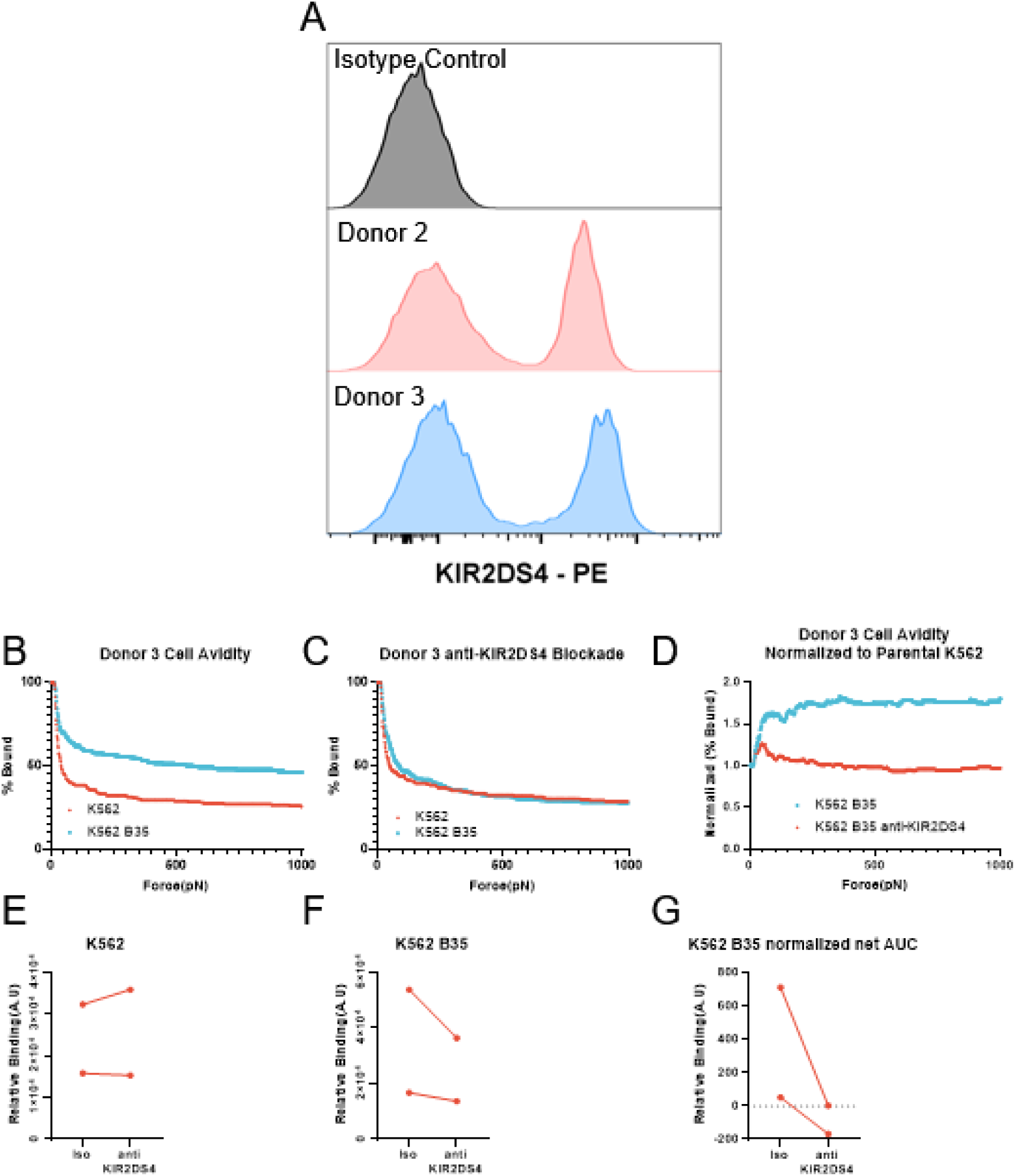
Donor KIR2DS4 typing and Cell Avidity Assay Example. (A) Example flow cytometry histogram of KIR2DS4-PE immunolabeling indicating Donor’s 2 and 3 positive protein expression. (B) Donor 3 single NK cells avidity versus K562 and K562 B*35:01 mono-allelic cells. (C) Anti-KIR2DS4 blockade. (D) Normalized graph to parental K562 cells. (E) Relative change in binding due to KIR2DS4 blockade of parental K562 cells for Donor 2, 3. (F) Relative change in binding due to KIR2DS4 blockade of K562 B*35:01 mono-allelic cells for Donor 2, 3. (G) Area under the curve (AUC) of normalized plots indicating degree of binding inhibition from KIR2DS4 blockade for Donor 2 and 3.

**Figure S3:**
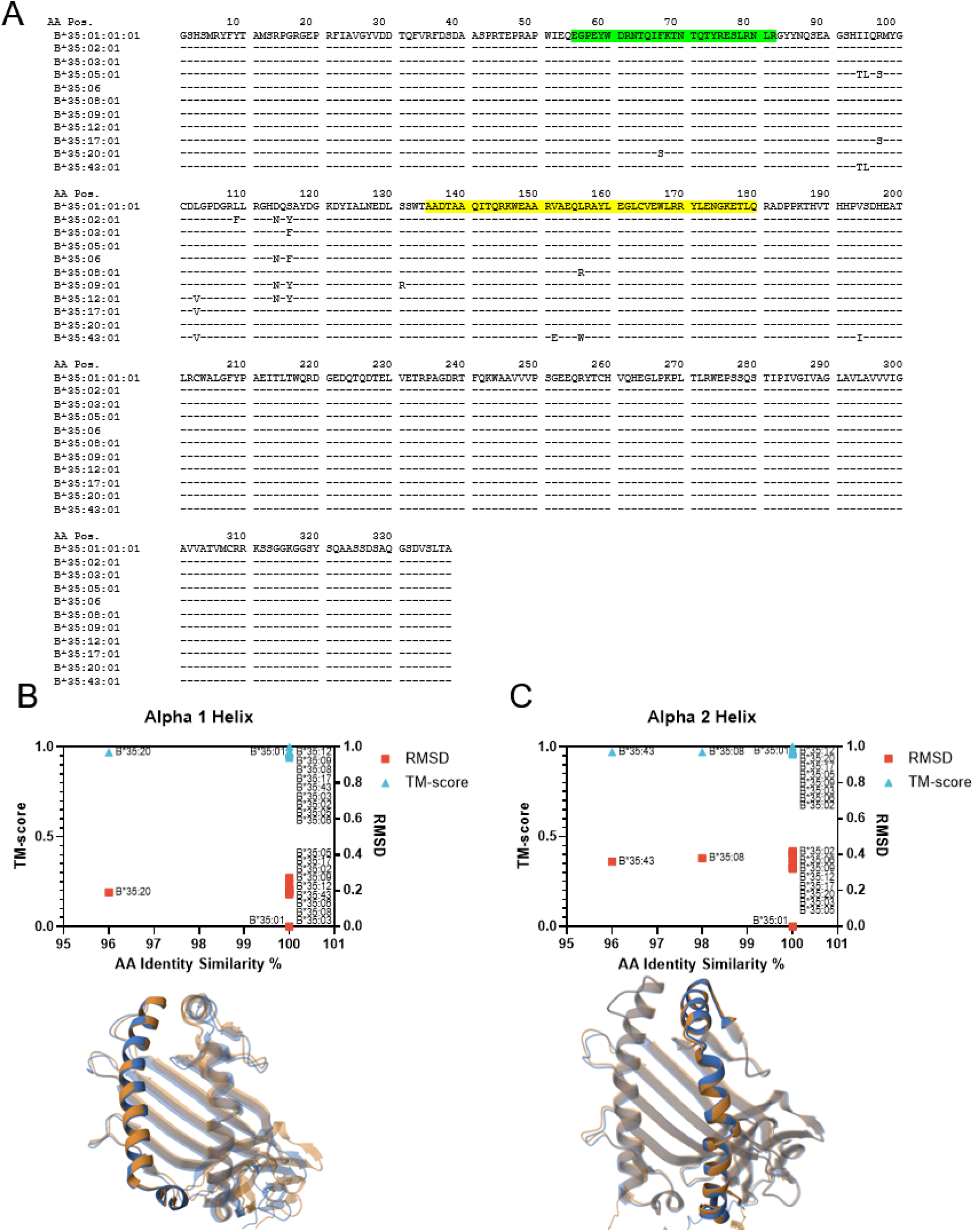
Breadth of HLA-B*35 allotype protein sequences expressed by patients in our study. (A) Mature protein sequence alignment with highlighted alpha helix 1 in green and alpha helix 2 in yellow. Changes are indicated by new amino acid single letter code. (B) Alphafolded3 folded protein structures compared to known HLA-B*35:01 crystal structure and then surface and backbone carbon chain aligned in PDB. RMSD and TM-Score plotted where RMSD=0 is perfect chain alignment and TM-score=1 is perfect surface alignment. Alpha helix 1 shown with a single overlay. (C) RMSD and TM-score plot for Alpha helix 2 shown with a single PDB overlay shown.

**Figure S4:**
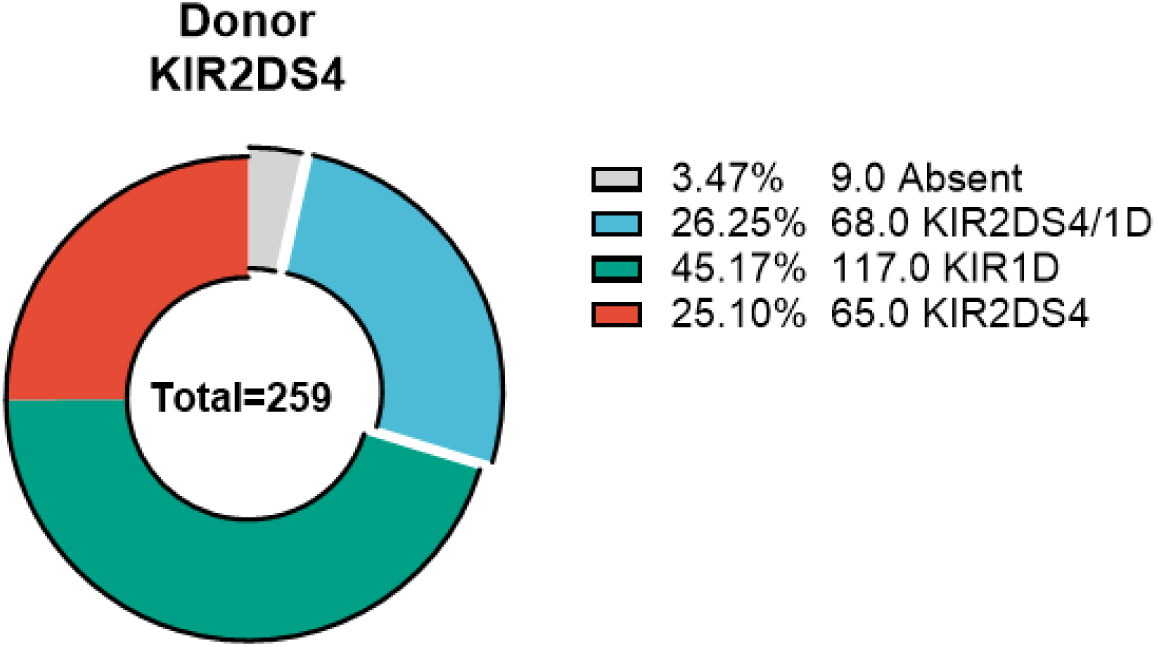
Total tested donor pool KIR2DS4 expression. Donut plot detailing the functional expression rates of each KIR2DS4 combination. KIR2DS4 indicates only full length, functional, KIR2DS4 detected, KIR2DS4/1D indicates both full length and deleted, indicates only KIR1D variant detected, Absent indicates no detection of either full length or deleted variant genes.

**Figure S5:**
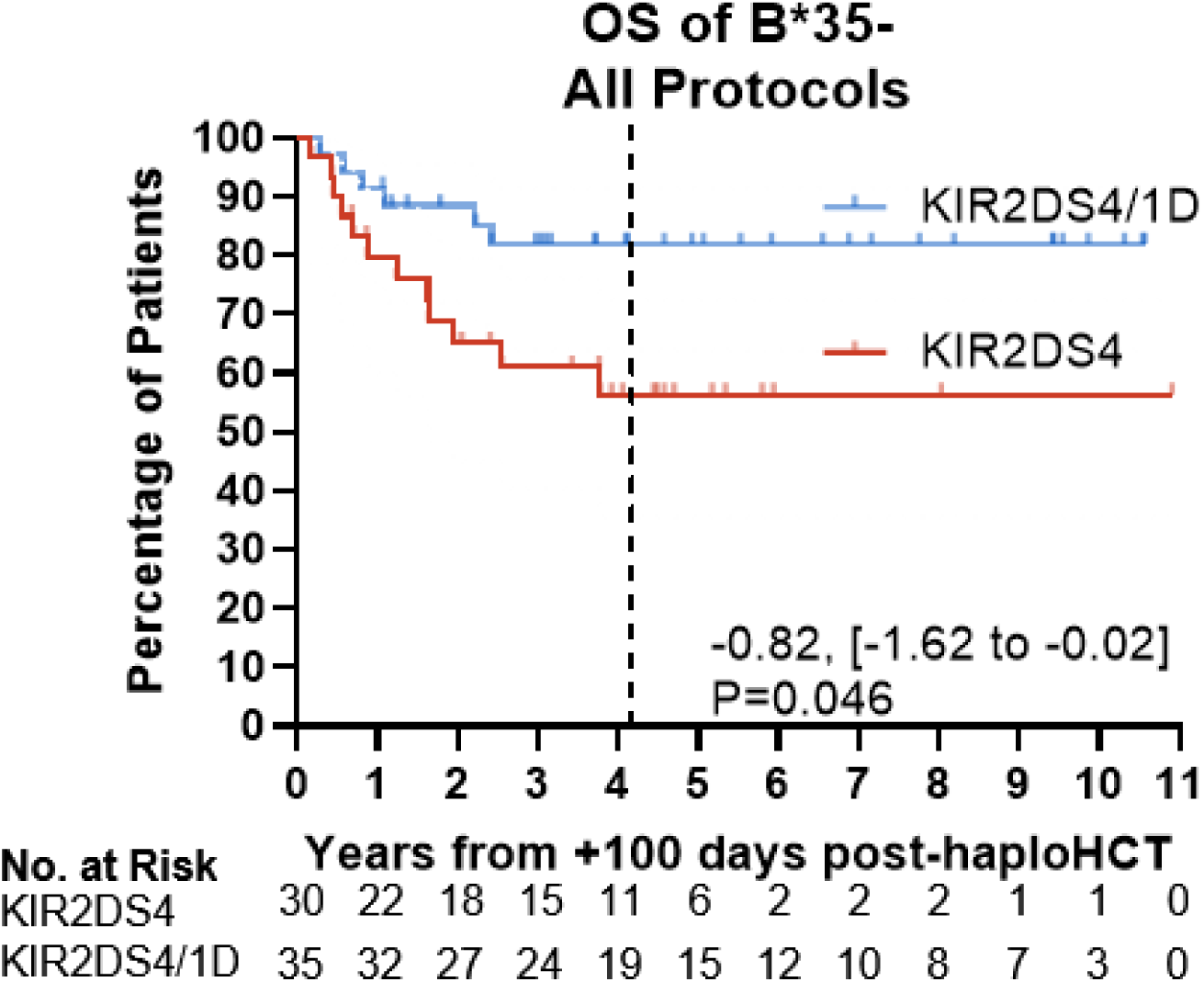
HLA-B*35 negative patients stratified by Donor KIR2DS4 expression. Event-free patients at day 100 post-haploHCT were separated based on their HLA-B*35 status and then subset by KIR2DS4 full length only (KIR2DS4) or full length with deleted variant (KIR2DS4/1D). Difference in 5-year RMST (years) is indicated with change from reference KIR2DS4/1D indicated with 95% confidence interval upper and lower bounds in brackets and two-sided Wald-based p-value. The number of patients at risk over the years is indicated below each graph. Kaplan-Meier survival curve of HLA-B*35 negative patients with KIR2DS4 and KIR2DS4/1D stratified donors.

**Figure S6:**
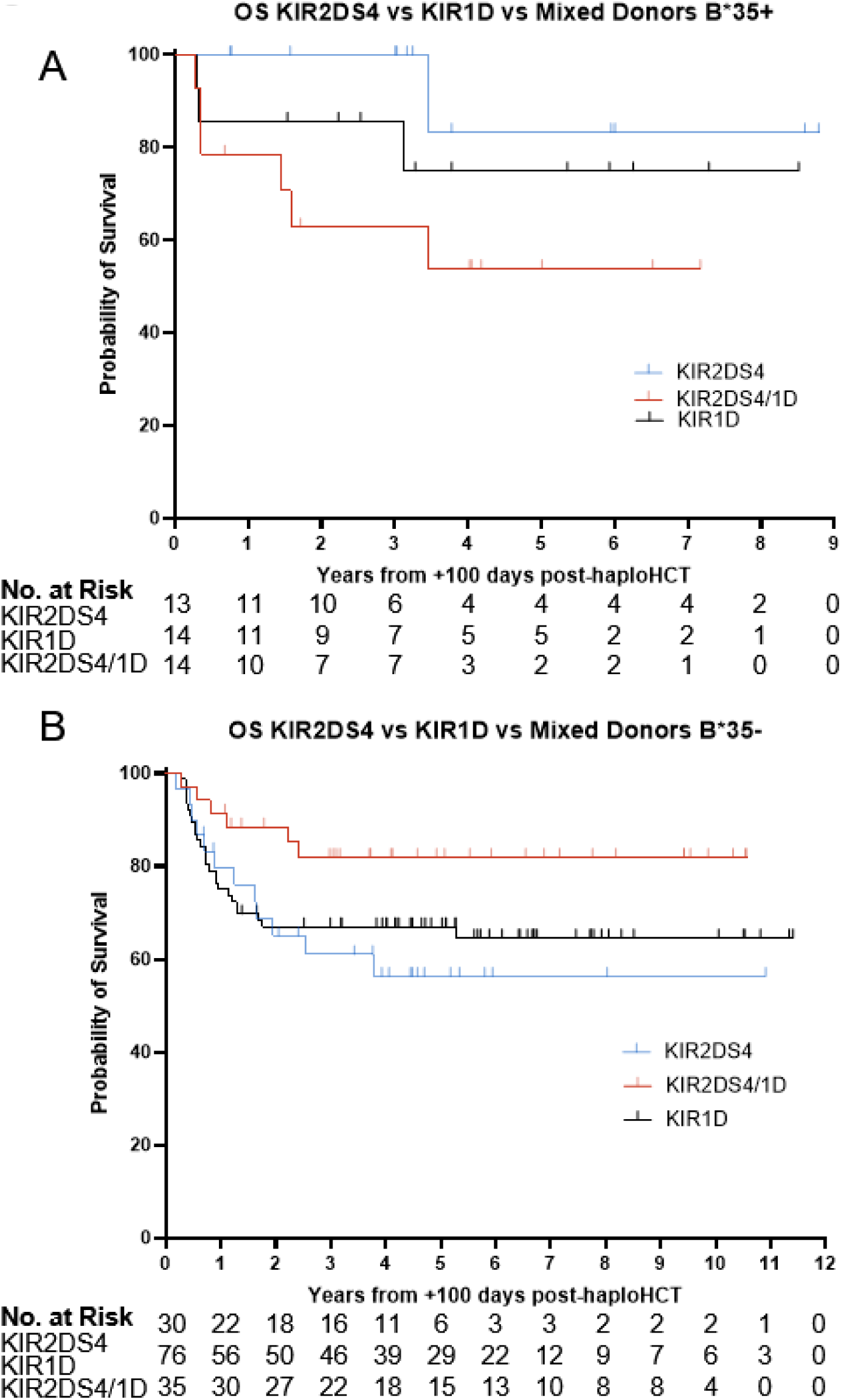
Pediatric patients treated with KIR2DS4, KIR1D, or Mixed donors. Event-free patients at day 100 post-haploHCT were separated based on their HLA-B*35 status and then subset by KIR2DS4 full length only (KIR2DS4) or full length with deleted variant (KIR2DS4/1D), or purely KIR1D. (A) HLA-B*35 positive patients. (B) HLA-B*35 negative patients.

**Table S1.** P-values for testing the difference in RMSTs for each analysis population and endpoint (rows) across varying time horizons (columns)

| Cohort | Endpoint | Year1 | Year2 | Year3 | Year4 | Year5 (Main) | Max |
| --- | --- | --- | --- | --- | --- | --- | --- |
| **B\*35+** | OS | 0.081 | 0.073 | 0.085 | 0.056 | 0.061 | 0.097 |
|  | RFS | 0.172 | 0.163 | 0.081 | 0.063 | 0.055 | 0.055 |
|  | OS, HAPNK | 0.264 | 0.111 | 0.081 | 0.030 | 0.016 | 0.007 |
|  | RFS, HAPNK | 0.155 | 0.053 | 0.005 | 0.002 | 0.001 | 0.001 |
| **B\*35-** | OS | 0.297 | 0.168 | 0.076 | 0.063 | 0.046 | 0.032 |
|  | RFS | 0.032 | 0.050 | 0.052 | 0.052 | 0.034 | 0.034 |
|  | OS, HAPNK | 0.785 | 0.649 | 0.459 | 0.322 | 0.205 | 0.095 |
|  | RFS, HAPNK | 0.200 | 0.257 | 0.286 | 0.237 | 0.142 | 0.142 |

In Table S1, we see that there is sensitivity in the p-values to very early time horizons (especially year 1 post-haploHCT) with lower strength of differences in RMST, but results are qualitatively similar at later time horizons. The direction of association (not reported) does not change with varying time horizon. The longest follow-up time for RFS is 5 years, so there is no difference between the 5-year RMSTs and “max” RMSTs for RFS.

**Table S2.** Results for differences in 5-year RMST adjusting for disease status and CMV status*.

| Cohort | Endpoint | Difference in 5-year RMST<br>(reference: KIR2DS4/1D) | 95% LB | 95% UB | P-Value |
| --- | --- | --- | --- | --- | --- |
| **B\*35+** | OS | 1.27 | -0.04 | 2.57 | 0.057 |
|  | RFS | 1.96 | 0.52 | 3.40 | 0.008 |
| **B\*35-** | OS | -0.79 | -1.59 | 0.01 | 0.054 |
|  | RFS | -0.91 | -1.80 | -0.03 | 0.044 |
|  | OS, HAPNK | -0.78 | -2.03 | 0.46 | 0.218 |
|  | RFS, HAPNK | -1.05 | -2.47 | 0.38 | 0.150 |

Results in Table S2 suggest that the main text unadjusted results do not appear importantly sensitive to adjustment by disease status or CMV status. We see that Table S2 point estimates and p-values are generally similar to corresponding main text results. The result for the B\*35+ cohort and RFS endpoint has the same direction and stronger evidence of association after adjustment. Importantly, due to the limited sample sizes, we could not perform adjusted 5-year RMST analyses for the HAPNK subgroup of the B\*35+ cohort with the RMST approach. Below, we used multivariable firthCox models that do fit for this subgroup.
**\*No RMST adjusted analysis could be performed for the B\*35+ HAPNK subgroup for OS or RFS endpoints.**

**Table S3.** Results for differences in 5-year Kaplan-Meier (KM) estimates.

| Cohort | Endpoint | KIR2DS<br>4/1D KM<br>estimate | KIR2DS<br>4 KM<br>estimate | Difference in 5-year<br>KM (reference:<br>KIR2DS4/1D) | 95% LB | 95% UB | P-Value |
| --- | --- | --- | --- | --- | --- | --- | --- |
| **B\*35+** | OS | 0.47 | 0.75 | 0.29 | -0.10 | 0.68 | 0.152 |
|  | RFS | 0.45 | 0.83 | 0.38 | 0.02 | 0.74 | 0.040 |
|  | OS,<br>HAPNK | 0.40 | 1.00 | 0.60 | 0.17 | 1.03 | 0.006 |
|  | RFS,<br>HAPNK | 0.25 | 1.00 | 0.75 | 0.33 | 1.17 | 0.001 |
| **B\*35-** | OS | 0.82 | 0.56 | -0.26 | -0.49 | -0.03 | 0.030 |
|  | RFS | 0.79 | 0.48 | -0.32 | -0.56 | -0.07 | 0.011 |
|  | OS,<br>HAPNK | 0.85 | 0.50 | -0.35 | -0.71 | 0.02 | 0.064 |
|  | RFS,<br>HAPNK | 0.85 | 0.40 | -0.45 | -0.81 | -0.08 | 0.016 |

Table S3 reports the supplementary analysis using 5-year KM estimates. Relative to the 5-year RMST analysis, we see that the B\*35+ OS endpoint has a weaker estimated effect size in the KM analysis but stronger or similar estimated effect sizes for RFS and for OS and RFS in the HAPNK subgroup. The B\*35- analyses have the same directions of effect and stronger evidence of association than the 5-year RMST analysis.

**Table S4.** Results for conventional Cox model.

| Cohort | Endpoint | HR | 95% LB** | 95% UB** | LRT p | Score P-Value |
| --- | --- | --- | --- | --- | --- | --- |
| **B\*35+** | OS | 0.35 | 0.09 | 1.40 | 0.124 | 0.121 |
|  | RFS | 0.25 | 0.05 | 1.26 | 0.068 | 0.070 |
|  | OS, HAPNK* |  |  |  | 0.026 | 0.049 |
|  | RFS, HAPNK* |  |  |  | 0.014 | 0.022 |
| **B\*35-** | OS | 2.72 | 1.02 | 7.25 | 0.038 | 0.037 |
|  | RFS | 2.84 | 1.15 | 7.05 | 0.019 | 0.019 |
|  | OS, HAPNK | 3.69 | 0.71 | 19.04 | 0.097 | 0.095 |
|  | RFS, HAPNK | 4.70 | 0.94 | 23.42 | 0.039 | 0.038 |

\*LRT and score p-values could be computed, but due to the limited numbers of events, model fitting is likely inappropriate for the B\*35+ HAPNK subgroup
\*\*Confidence intervals are Wald-based and may not correspond to LRT or score p-values

**Table S5.**
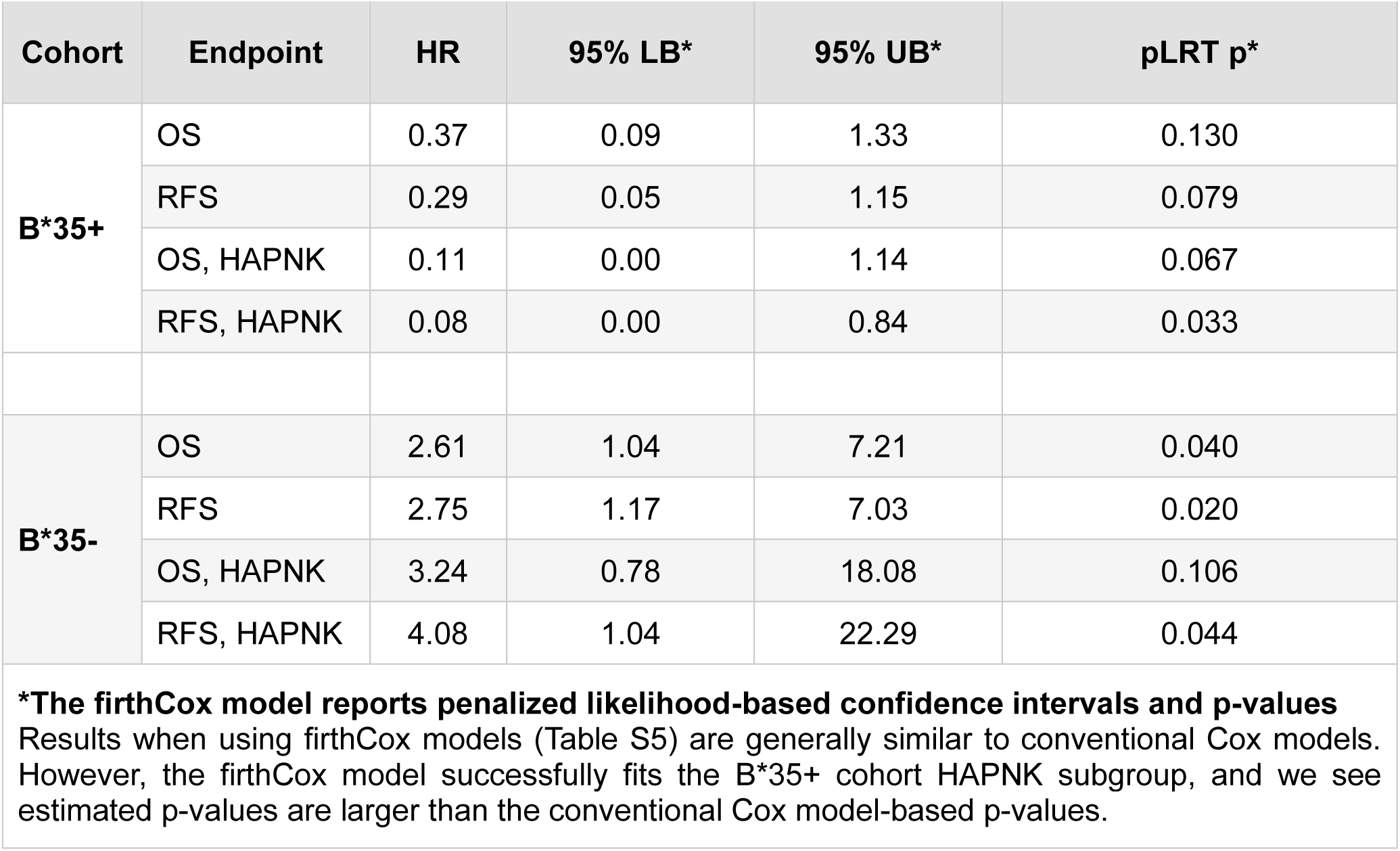
Results for unadjusted firthCox model.

| Cohort | Endpoint | HR | 95% LB* | 95% UB* | pLRT p* |
| --- | --- | --- | --- | --- | --- |
| **B\*35+** | OS | 0.37 | 0.09 | 1.33 | 0.130 |
|  | RFS | 0.29 | 0.05 | 1.15 | 0.079 |
|  | OS, HAPNK | 0.11 | 0.00 | 1.14 | 0.067 |
|  | RFS, HAPNK | 0.08 | 0.00 | 0.84 | 0.033 |
| **B\*35-** | OS | 2.61 | 1.04 | 7.21 | 0.040 |
|  | RFS | 2.75 | 1.17 | 7.03 | 0.020 |
|  | OS, HAPNK | 3.24 | 0.78 | 18.08 | 0.106 |
|  | RFS, HAPNK | 4.08 | 1.04 | 22.29 | 0.044 |

**\*The firthCox model reports penalized likelihood-based confidence intervals and p-values**
Results when using firthCox models (Table S5) are generally similar to conventional Cox models. However, the firthCox model successfully fits the B\*35+ cohort HAPNK subgroup, and we see estimated p-values are larger than the conventional Cox model-based p-values.

**Table S6.**
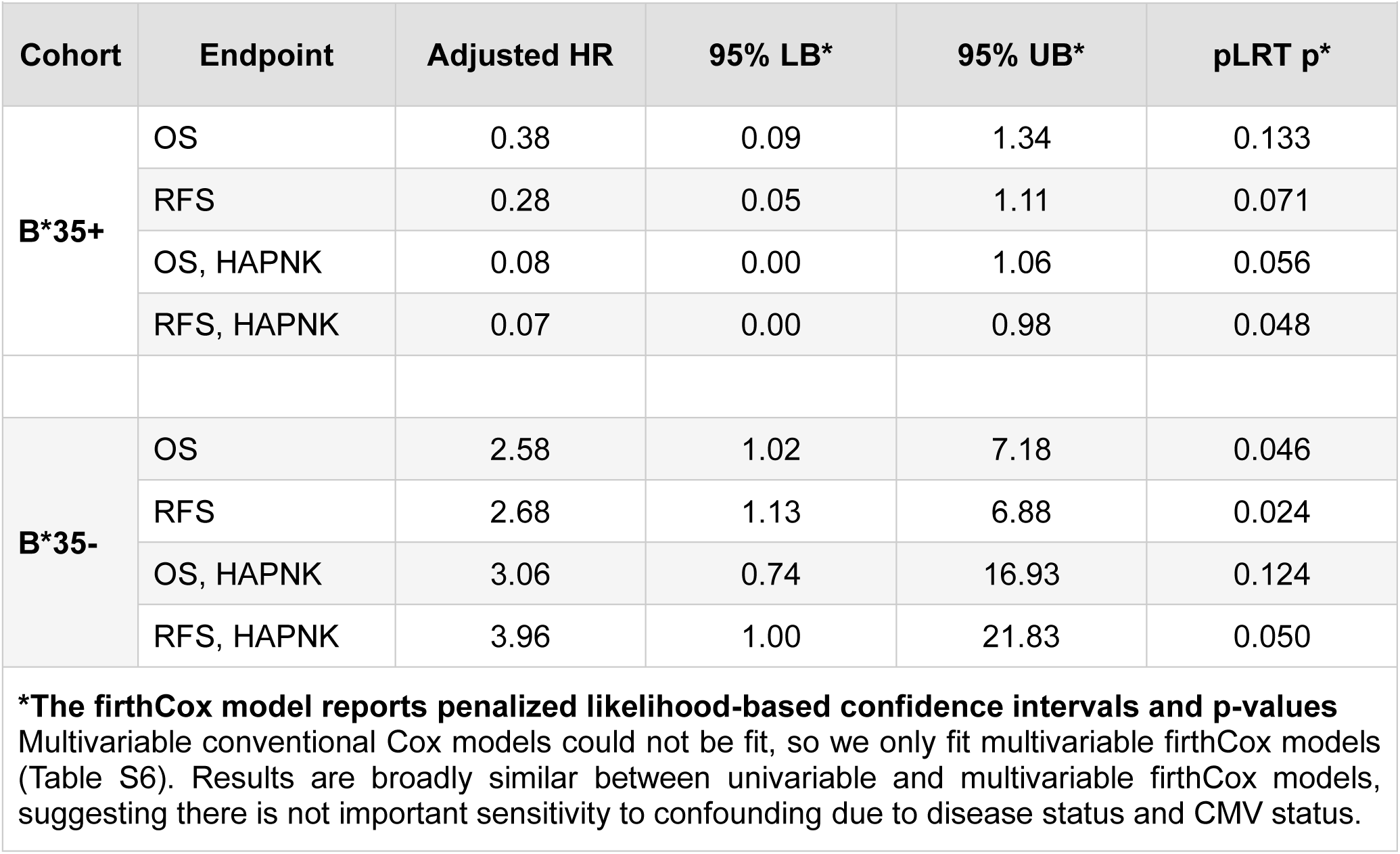
Results for firthCox model adjusting for disease status and CMV status.

| Cohort | Endpoint | Adjusted HR | 95% LB* | 95% UB* | pLRT p* |
| --- | --- | --- | --- | --- | --- |
| **B\*35+** | OS | 0.38 | 0.09 | 1.34 | 0.133 |
|  | RFS | 0.28 | 0.05 | 1.11 | 0.071 |
|  | OS, HAPNK | 0.08 | 0.00 | 1.06 | 0.056 |
|  | RFS, HAPNK | 0.07 | 0.00 | 0.98 | 0.048 |
| **B\*35-** | OS | 2.58 | 1.02 | 7.18 | 0.046 |
|  | RFS | 2.68 | 1.13 | 6.88 | 0.024 |
|  | OS, HAPNK | 3.06 | 0.74 | 16.93 | 0.124 |
|  | RFS, HAPNK | 3.96 | 1.00 | 21.83 | 0.050 |

**\*The firthCox model reports penalized likelihood-based confidence intervals and p-values**
Multivariable conventional Cox models could not be fit, so we only fit multivariable firthCox models (Table S6). Results are broadly similar between univariable and multivariable firthCox models, suggesting there is not important sensitivity to confounding due to disease status and CMV status.

**Table S7.** Results for differences in 5-year RMST using OS* and RFS* defined from date of HCT

| Cohort | Endpoint | KIR2DS4/<br>1D 5-year<br>RMST | KIR2DS4<br>5-year<br>RMST | Difference in<br>5-year RMST<br>(reference:<br>KIR2DS4/1D) | 95% LB | 95% UB | P-Value |
| --- | --- | --- | --- | --- | --- | --- | --- |
| **B\*35+** | OS* | 3.06 | 4.06 | 0.99 | -0.43 | 2.42 | 0.173 |
|  | RFS* | 2.56 | 3.70 | 1.13 | -0.48 | 2.75 | 0.169 |
|  | OS*, HAPNK | 2.95 | 4.21 | 1.26 | -0.93 | 3.45 | 0.259 |
|  | RFS*, HAPNK | 1.66 | 4.19 | 2.52 | 0.39 | 4.66 | 0.020 |
| **B\*35-** | OS* | 3.94 | 3.56 | -0.38 | -1.26 | 0.49 | 0.394 |
|  | RFS* | 3.80 | 3.25 | -0.55 | -1.50 | 0.40 | 0.258 |
|  | OS*, HAPNK | 3.83 | 3.58 | -0.25 | -1.68 | 1.17 | 0.727 |
|  | RFS*, HAPNK | 3.80 | 3.28 | -0.52 | -2.10 | 1.05 | 0.515 |

In Table S7 we report the analyses when not using the 100-day landmark analysis for the main text 5-year RMST analysis. We observe that directions of associations are the same, but the strengths of association are weaker. From corresponding KM curves (not reported), we see early events are more equally distributed between the two groups.

## Supplemental Appendix Protocol Table

| **Supplemental Appendix Protocol Table** |  |  |
| --- | --- | --- |
| **NCT Number** | **Clinical trial acronym** | **Number of patients** |
| NCT03849651 | HAP2HCT | 30 |
| NCT01621477 | HAP3R | 5 |
| NCT01807611 | HAPNK1 | 36 |
| NCT02442660 | HIFLEX | 1 |
| N/A | NPTP | 5 |
| NCT02790515 | REF2HCT | 16 |
| NCT02259348 | REFNK1 | 1 |
| N/A | SPIP* | 1 |

\*SPIP per REF2HCT

### Appendix: DRST members contributing to this study

Dr. Georg-Nikolaus Franke, Universitätsklinikum Leipzig, 18 patients Prof. Ahmet Elmaagacli, Asklepios Klinik St. Georg, 12 patients

Dr. Elisa Sala, Universitätsklinikum Ulm, 11 patients

Prof. Friedrich Stölzel, Universitätsklinikum Schleswig-Holstein / Campus Kiel, 10 patients Dr. Eva Wagner-Drouet, Universitätsmedizin Mainz, 10 patients

Prof. Gerald Wulf, Universitätsklinik, 7 patients

Dr. Johannes Lakner, Universitätsmedizin Rostock, 6 patients Dr. Daniel Teschner, Universitätsklinikum Würzburg, 6 patients

Dr. Jörg Thomas Bittenbring, Universitätsklinikum des Saarlandes, 6 patients Prof. Igor Wolfgang Blau, Charité - Universitätsmedizin Berlin, 6 patients Prof. Jochen Casper, Klinikum Oldenburg AöR, 4 patients

Dr. Martin Kaufmann, Robert-Bosch-Krankenhaus, 3 patients Dr. Arne Brecht, DKD HELIOS Klinik Wiesbaden, 2 patients

Dr. Stephan Kaun, Gesundheit Nord gGmbH, Klinikum Bremen-Mitte, 2 patients Dr. Kerstin Schäfer-Eckart, Klinikum Nürnberg Nord, 2 patients

Dr. Armin Gerbitz, Universitätsklinikum Erlangen, 2 patients

Prof. Mark Ringhoffer, Städt. Klinikum Karlsruhe gGmbH, 1 patient

Dr. Friederike Wortmann, Universitätsklinikum Schleswig-Holstein / Campus Lübeck, 1 patient

## References

1. Thomas ED, Lochte HL, Jr., Lu WC, Ferrebee JW. Intravenous infusion of bone marrow in patients receiving radiation and chemotherapy. N Engl J Med 1957;257(11):491–6. (In eng). DOI: 10.1056/NEJM195709122571102.

2. Timofeeva OA, Philogene MC, Zhang QJ. Current donor selection strategies for allogeneic hematopoietic cell transplantation. Hum Immunol 2022;83(10):674–686. (In eng). DOI: 10.1016/j.humimm.2022.08.007.

3. Pulsipher MA, Logan BR, Kiefer DM, et al. Related peripheral blood stem cell donors experience more severe symptoms and less complete recovery at one year compared to unrelated donors. Haematologica 2019;104(4):844–854. (In eng). DOI: 10.3324/haematol.2018.200121.

4. McCurdy SR, Zhang M-J, St. Martin A, et al. Effect of donor characteristics on haploidentical transplantation with posttransplantation cyclophosphamide. Blood Advances 2018;2(3):299–307. DOI: 10.1182/bloodadvances.2017014829.

5. Cusatis R, Litovich C, Feng Z, et al. Current Trends and Outcomes in Cellular Therapy Activity in the United States, Including Prospective Patient-Reported Outcomes Data Collection in the Center for International Blood and Marrow Transplant Research Registry. Transplant Cell Ther 2024;30(9):917.e1–917.e12. (In eng). DOI: 10.1016/j.jtct.2024.06.021.

6. O’Donnell PV, Jones RJ. The development of post-transplant cyclophosphamide: Half a century of translational team science. Blood Rev 2023;62:101034. (In eng). DOI: 10.1016/j.blre.2022.101034.

7. Petersen SL, Ryder LP, Björk P, et al. A comparison of T-, B- and NK-cell reconstitution following conventional or nonmyeloablative conditioning and transplantation with bone marrow or peripheral blood stem cells from human leucocyte antigen identical sibling donors. Bone Marrow Transplant 2003;32(1):65–72. (In eng). DOI: 10.1038/sj.bmt.1704084.

8. Thomson BG, Robertson KA, Gowan D, et al. Analysis of engraftment, graft-versus-host disease, and immune recovery following unrelated donor cord blood transplantation. Blood 2000;96(8):2703–11. (In eng).

9. Small TN, Papadopoulos EB, Boulad F, et al. Comparison of immune reconstitution after unrelated and related T-cell-depleted bone marrow transplantation: effect of patient age and donor leukocyte infusions. Blood 1999;93(2):467–80. (In eng).

10. Farag SS, Fehniger T, Ruggeri L, Velardi A, Caligiuri MA. Natural killer cells: biology and application in stem-cell transplantation. Cytotherapy 2002;4(5):445–446. DOI: 10.1080/146532402320776134.

11. Parham P, Moffett A. Variable NK cell receptors and their MHC class I ligands in immunity, reproduction and human evolution. Nat Rev Immunol 2013;13(2):133–44. (In eng). DOI: 10.1038/nri3370.

12. Moretta L, Moretta A. Killer immunoglobulin-like receptors. Curr Opin Immunol 2004;16(5):626–33. (In eng). DOI: 10.1016/j.coi.2004.07.010.

13. Ruggeri L, Capanni M, Urbani E, et al. Effectiveness of donor natural killer cell alloreactivity in mismatched hematopoietic transplants. Science 2002;295(5562):2097–100. DOI: 10.1126/science.1068440.

14. Igarashi T, Wynberg J, Srinivasan R, et al. Enhanced cytotoxicity of allogeneic NK cells with killer immunoglobulin-like receptor ligand incompatibility against melanoma and renal cell carcinoma cells. Blood 2004;104(1):170–7. (In eng). DOI: 10.1182/blood-2003-12-4438.

15. Ruggeri L, Capanni M, Casucci M, et al. Role of natural killer cell alloreactivity in HLA-mismatched hematopoietic stem cell transplantation. Blood 1999;94(1):333–9. (In eng).

16. Schetelig J, Baldauf H, Heidenreich F, et al. Donor KIR genotype based outcome prediction after allogeneic stem cell transplantation: no land in sight. Front Immunol 2024;15:1350470. (In eng). DOI: 10.3389/fimmu.2024.1350470.

17. Fein JA, Shouval R, Krieger E, et al. Systematic evaluation of donor-KIR/recipient-HLA interactions in HLA-matched hematopoietic cell transplantation for AML. Blood Adv 2024;8(3):581–590. DOI: 10.1182/bloodadvances.2023011622.

18. Mehta RS, Rezvani K. Can we make a better match or mismatch with KIR genotyping? Hematology Am Soc Hematol Educ Program 2016;2016(1):106–118. (In eng). DOI: 10.1182/asheducation-2016.1.106.

19. Gowdavally S, Tsamadou C, Platzbecker U, et al. KIR2DS4 and Its Variant KIR1D in KIR-AA Genotype Donors Showed Differential Survival Impact in Patients with Lymphoid Disease after HLA-Matched Unrelated Hematopoietic Stem Cell Transplantation. Transplant Cell Ther 2023;29(7):457 e1–457 e10. DOI: 10.1016/j.jtct.2023.04.006.

20. Middleton D, Gonzalez A, Gilmore PM. Studies on the Expression of the Deleted KIR2DS4*003 Gene Product and Distribution of KIR2DS4 Deleted and Nondeleted Versions in Different Populations. Human Immunology 2007;68(2):128–134. DOI: 10.1016/j.humimm.2006.12.007.

21. Wu X, Yao Y, Bao X, et al. KIR2DS4 and Its Variant KIR1D Are Associated with Acute Graft-versus-Host Disease, Cytomegalovirus, and Overall Survival after Sibling-Related HLA-Matched Transplantation in Patients with Donors with KIR Gene Haplotype A. Biol Blood Marrow Transplant 2016;22(2):220–225. DOI: 10.1016/j.bbmt.2015.10.004.

22. Bao X, Hou L, Sun A, Chen M, Chen Z, He J. An allelic typing method for 2DS4 variant used in study of haplotypes of killer cell immunoglobulin-like receptor gene. Int J Lab Hematol 2010;32(6 Pt 2):625–32. (In eng). DOI: 10.1111/j.1751-553X.2010.01234.x.

23. Abramson J, Adler J, Dunger J, et al. Accurate structure prediction of biomolecular interactions with AlphaFold 3. Nature 2024;630(8016):493–500. DOI: 10.1038/s41586-024-07487-w.

24. Barker DJ, Maccari G, Georgiou X, et al. The IPD-IMGT/HLA Database. Nucleic Acids Res 2023;51(D1):D1053–d1060. (In eng). DOI: 10.1093/nar/gkac1011.

25. Berman HM, Westbrook J, Feng Z, et al. The Protein Data Bank. Nucleic Acids Research 2000;28(1):235–242. DOI: 10.1093/nar/28.1.235.

26. Yanaka S, Ueno T, Shi Y, et al. Peptide-dependent Conformational Fluctuation Determines the Stability of the Human Leukocyte Antigen Class I Complex*. Journal of Biological Chemistry 2014;289(35):24680–24690. DOI: 10.1074/jbc.M114.566174.

27. Chockley P, Patil SL, Gottschalk S. Transient blockade of TBK1/IKKepsilon allows efficient transduction of primary human natural killer cells with vesicular stomatitis virus G-pseudotyped lentiviral vectors. Cytotherapy 2021;23(9):787–792. (In eng). DOI: 10.1016/j.jcyt.2021.04.010.

28. Pesce S, Carlomagno S, Moretta A, Sivori S, Marcenaro E. Uptake of CCR7 by KIR2DS4(+) NK cells is induced upon recognition of certain HLA-C alleles. J Immunol Res 2015;2015:754373. DOI: 10.1155/2015/754373.

29. Graef T, Moesta AK, Norman PJ, et al. KIR2DS4 is a product of gene conversion with KIR3DL2 that introduced specificity for HLA-A*11 while diminishing avidity for HLA-C. J Exp Med 2009;206(11):2557–72. (In eng). DOI: 10.1084/jem.20091010.

30. Zhou Y, Smith J, Keerthi D, et al. Longitudinal clinical data improve survival prediction after hematopoietic cell transplantation using machine learning. Blood Adv 2024;8(3):686–698. DOI: 10.1182/bloodadvances.2023011752.

31. Mamcarz E, Madden R, Qudeimat A, et al. Improved survival rate in T-cell depleted haploidentical hematopoietic cell transplantation over the last 15 years at a single institution. Bone Marrow Transplant 2020;55(5):929–938. DOI: 10.1038/s41409-019-0750-7.

32. Anderson JR, Davis RB. Analysis of survival by tumor response. J Clin Oncol 1986;4(1):115–7. (In eng). DOI: 10.1200/jco.1986.4.1.115.

33. Chockley PJ, Ibanez-Vega J, Krenciute G, Talbot LJ, Gottschalk S. Synapse-tuned CARs enhance immune cell anti-tumor activity. Nat Biotechnol 2023;41(10):1434–1445. (In eng). DOI: 10.1038/s41587-022-01650-2.

34. Naik S, Li Y, Talleur AC, et al. Memory T-cell enriched haploidentical transplantation with NK cell addback results in promising long-term outcomes: a phase II trial. J Hematol Oncol 2024;17(1):50. (In eng). DOI: 10.1186/s13045-024-01567-0.

35. Single RM, Martin MP, Gao X, et al. Global diversity and evidence for coevolution of KIR and HLA. Nat Genet 2007;39(9):1114–9. (In eng). DOI: 10.1038/ng2077.

36. Hilton HG, Moesta AK, Guethlein LA, Blokhuis J, Parham P, Norman PJ. The production of KIR-Fc fusion proteins and their use in a multiplex HLA class I binding assay. J Immunol Methods 2015;425:79–87. (In eng). DOI: 10.1016/j.jim.2015.06.012.

37. Chaturantabut S, Oliver S, Frederick DT, et al. Identification of potent biparatopic antibodies targeting FGFR2 fusion-driven cholangiocarcinoma. J Clin Invest 2025;135(8) (In eng). DOI: 10.1172/jci182417.

38. Orange JS. Formation and function of the lytic NK-cell immunological synapse. Nat Rev Immunol 2008;8(9):713–25. (In eng). DOI: 10.1038/nri2381.

39. Kamsma D, Bochet P, Oswald F, et al. Single-Cell Acoustic Force Spectroscopy: Resolving Kinetics and Strength of T Cell Adhesion to Fibronectin. Cell Reports 2018;24(11):3008–3016. DOI: 10.1016/j.celrep.2018.08.034.

40. Heinze G, Schemper M. A solution to the problem of monotone likelihood in Cox regression. Biometrics 2001;57(1):114–9. (In eng). DOI: 10.1111/j.0006-341x.2001.00114.x.

